# The long-term impacts of domestic and international TB service improvements on TB trends within the United States: a mathematical modelling study

**DOI:** 10.1101/2024.03.20.24304585

**Authors:** Nicolas A Menzies, Nicole A Swartwood, Ted Cohen, Suzanne M Marks, Susan A Maloney, Courtney Chappelle, Jeffrey W Miller, Garrett R Beeler Asay, Anand A Date, C Robert Horsburgh, Joshua A Salomon

**Author notes:** **Corresponding Author:** Nicolas A. Menzies, Department of Global Health and Population, Harvard T.H. Chan School of Public Health, 665 Huntington Ave, Boston, MA 02115, USA. **Disclaimer**: The findings and conclusions in this report are those of the authors and do not necessarily represent the views of the Centers for Disease Control and Prevention.

## Abstract

**Background:** For low TB incidence settings, disease elimination is a long-term goal. We investigated pathways to TB pre-elimination and elimination (incidence <1.0 and <0.1 per 100,000, respectively) in the United States.

**Methods:** Using a mathematical modelling framework, we simulated how U.S. TB incidence could be affected by changes in TB services in the countries of origin for future migrants to the United States, as well as changes in TB services inside the United States. We constructed intervention scenarios representing improvements in TB services internationally and within the United States, individually and in combination, plus a base-case scenario representing continuation of current services. We simulated health and economic outcomes until 2100.

**Findings:** Under the base-case, U.S. TB incidence rates were projected to decline to 1.8 (95% uncertainty interval: 1.5, 2.1) per 100,000 by 2050. Intervention scenarios produced substantial reductions in TB incidence, with the combination of all domestic and international interventions projected to achieve pre-elimination by 2033 (2031, 2037). Compared to the base-case, this combination could avert 101 (84, 120) thousand U.S. TB cases and 13 (11, 16) thousand U.S. TB deaths over 2025-2050; the total economic value of these TB incidence reductions was estimated as US$68 (33, 112) billion. TB elimination was not projected before 2100.

**Interpretation:** Strengthening TB services domestically, promoting the development of more effective technologies and interventions, and supporting TB programs in high-burden countries are key strategies for accelerating progress towards TB elimination in the United States.

**Funding:** U.S. Centers for Disease Control and Prevention.

**Research in Context:** *Evidence before this study:* A number of studies have investigated the potential health impacts of tuberculosis intervention options in individual countries, across high incidence and low incidence settings. Studies in high incidence settings have generally emphasized TB case detection as a high-impact strategy, while studies in low incidence settings have focused on preventive treatment among individuals with TB infection. Few studies have examined the combined effect of interventions choices in both high- and low-incidence settings, and how these choices can contribute to progress towards TB elimination goals in individual countries.

*Added value of this study:* Using a mathematical modelling approach, this study assessed how interventions in high burden countries and interventions used within the United States could affect future TB trends within the United States. Dependencies between different TB epidemics were modelled explicitly through migration. The analysis examined the extent to which different intervention combinations could accelerate progress towards TB pre-elimination and elimination goals (incidence <1.0 and <0.1 per 100,000, respectively).

*Implications of all the available evidence:* In this low-burden setting, actions to strengthen TB services domestically, promote the development of more effective technologies and interventions, and support TB programs in high-burden countries were all found to be complementary and impactful strategies for accelerating progress towards TB elimination.

## Introduction

Eliminating TB is the aspirational goal of global TB policy, with aggressive action to combat the disease attracting high-level political commitments,^1,2^ and with targets towards this goal included in the WHO End TB Strategy and Sustainable Development Goals.^3,4^ While global efforts to reduce TB incidence have accelerated slowly^5^—and been delayed by the COVID-19 pandemic^6^—technological advances are providing new tools for TB prevention and care.^7–10^ Despite setbacks, global TB strategy continues to target rapid progress.^11^

In low-incidence countries such as the United States, TB epidemiology is characterized by limited transmission concentrated within populations at greatest risk, with most TB cases resulting from progression of established *Mycobacterium tuberculosis* (*Mtb*) infections acquired in past years. In many of these countries, a substantial fraction of TB cases arises among migrants from high TB burden countries, due to infections acquired before migration.^12^ In the United States only 12% of genotyped TB cases during 2020–2021 were estimated to result from recent transmission, and 71% of all TB cases in 2021 arose in the non-U.S.-born population.^13^ For countries like the United States, strategies for pursuing TB elimination include expanded testing and preventive treatment for *Mtb* infection among individuals at increased risk of TB disease (including persons born in countries with high TB incidence and other populations with elevated *Mtb* infection prevalence or medical risk factors for rapid disease progression),^14,15^ rapid detection of new TB cases and contact investigation to interrupt transmission, strengthening care cascades for individuals with *Mtb* infection and TB disease, encouraging new technologies, and engagement in global TB prevention and care effort.^16,17^

While elimination is a long-established goal of U.S. TB policy,^18,19^ current TB incidence— estimated at 2.5 per 100,000 for 2022^20^—is well above established thresholds for pre-elimination and elimination, defined as TB incidence of <1.0 per 100,000 and <0.1 per 100,000, respectively.^21^ A long historical decline in U.S. incidence rates has flattened in recent years,^13^ while since 2017 non-U.S.-born persons have represented over 70% of all U.S. TB cases.^13^ The gap in TB incidence rates between non-U.S.-born residents’ countries of birth and the United States has risen over time,^22^ and demographic projections suggest that non-U.S.-born persons will form an increasing share of the future U.S. population.^23^ For these reasons, it is likely that future U.S. TB trends will depend on the relative success of TB programs in other countries.^24,25^

In this study we examined possible intervention pathways towards TB elimination in the United States. Using mathematical modelling, we investigated approaches for strengthening TB services, both within and outside of the United States. For each approach we projected future TB incidence trends within the United States, the long-term health and economic outcomes of TB, and the timing of reaching pre-elimination and elimination thresholds.

## Methods

We constructed intervention scenarios representing improvements in TB services internationally and within the United States, as well as a base-case scenario representing continuation of current service coverage and effectiveness. Using mathematical modeling, we simulated future epidemiological outcomes to understand how each scenario might affect future TB incidence trends within the United States, and the number of years until pre-elimination (<1.0 TB case per 100,000) and elimination (<0.1 TB cases per 100,000) could be reached. We calculated additional outcomes quantifying health and economic benefits realized under each intervention scenario compared to the base-case. This study was determined to be non–human subjects research by the Harvard University Institutional Review Board (IRB22-1328).

### Mathematical model

We adapted a transmission-dynamic model of TB epidemiology, risk factors, and health services in the U.S. population (‘U.S. model’), previously used to study improvements in TB prevention, diagnosis, and treatment within the United States.^26–29^ Parameters describing TB natural history and U.S. TB services were based on published literature and routine reporting data.^13,29^ Data on the size of the non-U.S.-born population were drawn from the American Community Survey for 2000-2021,^30^ and used to estimate rates of migration into the United States by age, year, and birth country.^31^ We projected future migration trends to match U.S. Census Bureau projections.^23^ To estimate *Mtb* infection prevalence among current and future migrants, we created simplified TB models (‘non-U.S. models’) for each of the top 30 countries of birth by reported TB cases among non-U.S.-born individuals entering the United States after 2010 (Table S1), following a published approach.^24^ An additional model was created to represent remaining low- and middle-income countries not modelled individually, and another to represent remaining high-income countries. Figure S1 shows modelled health states and transitions.

We fit models to data for each setting using a Bayesian calibration approach.^32,33^ Non-U.S. models were calibrated to WHO-estimated TB incidence trends (Figure S2) and published *Mtb* infection prevalence estimates (Figure S3).^34,35^ The U.S. model was calibrated to Interferon-Gamma Release Assay (IGRA) data collected in the 2011-2012 round of the National Health and Nutrition Examination Survey (NHANES), which provide the most recent nationally-representative U.S. estimates of *Mtb* infection prevalence by age and U.S.-born status.^36^ This model was also calibrated to trends in reported TB cases within multiple population strata, extracted from the National TB Surveillance System.^37^ To ensure that TB trends among entering migrants matched available data, we calibrated the model to reproduce the number of TB cases among non-U.S.-born persons residing in the United States from each of the countries and country groups included in the non-U.S. models between 2000 and 2021 (Figures S4-S6).

### Analytic scenarios

We created a base-case scenario representing the continuation of current service coverage and effectiveness in each modelled setting. For non-U.S. models, we extrapolated future TB epidemiology based on TB incidence trends for each country over 2011-2021.^34^ For the United States we assumed the coverage and effectiveness of TB services would remain at current levels. We created hypothetical scenarios representing improvements in TB services in non-U.S. countries (‘international intervention scenarios’), based on the Stop TB Partnership’s Global Plan to End TB 2023-2030.^11^ These scenarios describe the service expansion and technological advances needed to achieve the WHO End TB Strategy targets (by 2035, a 95% reduction in TB mortality and 90% reduction in TB incidence rates compared to 2015).^3^ These scenarios were simulated for each of the 32 countries and country groups included in the non-U.S. models, matching the projections of the Stop TB Partnership’s Global Plan to End TB.^11^ Figure S7 shows incidence projections for each modelled setting under the international intervention scenarios. We created an additional set of scenarios representing improvements in TB services provided within or at the time of entry to the United States (‘domestic intervention scenarios’), based on an earlier modelling study.^29^ We created a third set of scenarios representing the combination of international and domestic service improvements. Scenarios are described in Table 1 (details in Supplement). As a sensitivity analysis we adjusted the intervention parameters of the U.S. model to achieve the changes targeted in the End TB Strategy (95% reduction in TB mortality and 90% reduction in TB incidence rates between 2015 and 2035), while TB trends in other countries were assumed to follow their current trajectories. Using the fitted mathematical models we simulated future U.S. TB outcomes for each scenario.

**Table 1:**
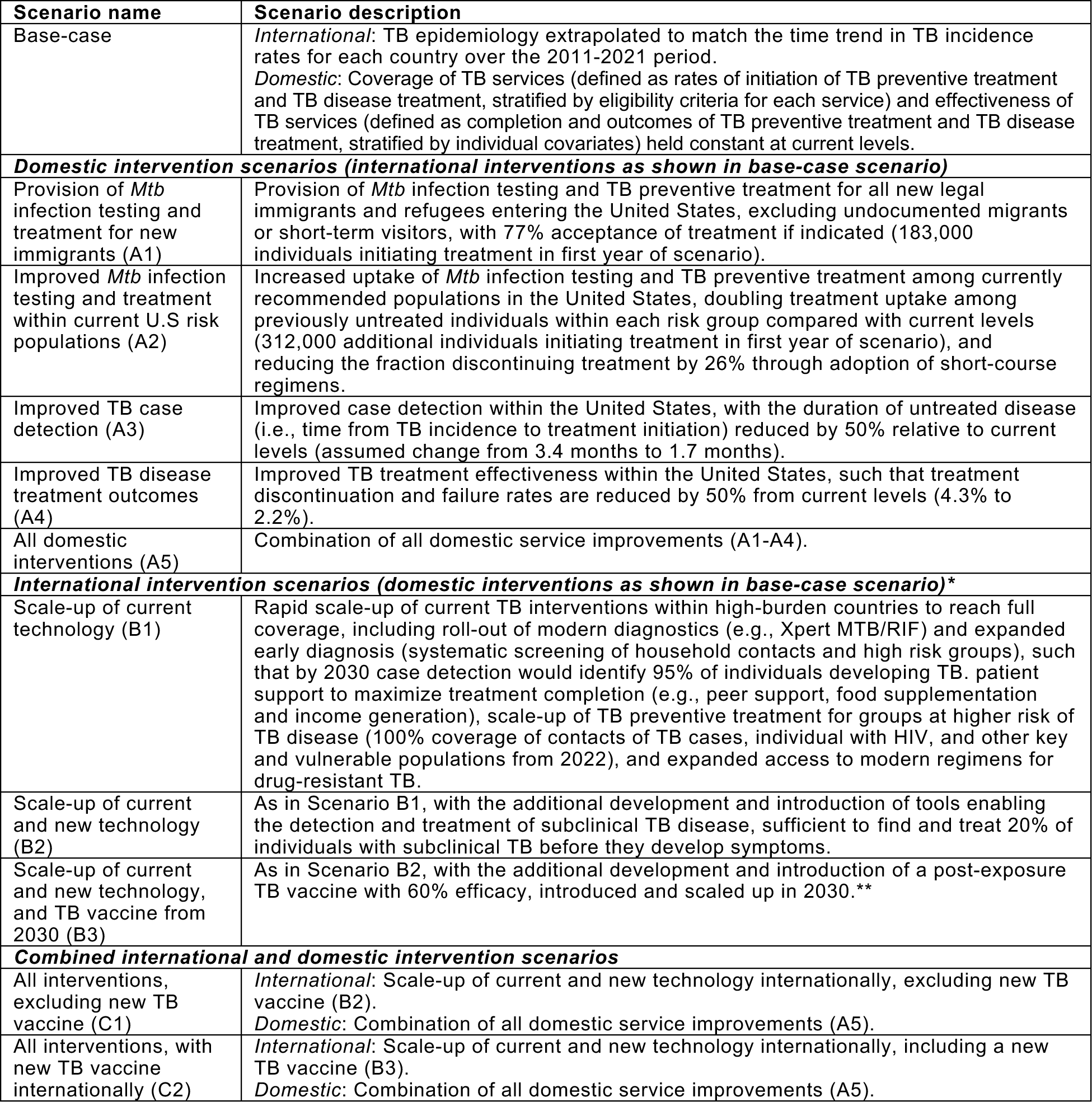
Base-case and intervention scenarios. * International scenarios are based on strategies included in the Global Plan to End TB ^11^, which represents the scale up of existing and anticipated TB interventions required to meet the WHO End TB Strategy targets, including a 95% reduction in TB deaths and 90% reduction in TB incidence rates between 2015 and 2035 ^3^. ** The Global Plan to End TB assumes that a vaccine will be introduced in 2025. However, given the progress on vaccine development at the time of this study it was decided that 2030 was a more plausible date for availability of a new TB vaccine. Additional details on the specification of analytic scenarios are given in the Supplement.

| Scenario name | Scenario description |
| --- | --- |
| Base-case | *International:* TB epidemiology extrapolated to match the time trend in TB incidence rates for each country over the 2011-2021 period.<br>*Domestic:* Coverage of TB services (defined as rates of initiation of TB preventive treatment and TB disease treatment, stratified by eligibility criteria for each service) and effectiveness of TB services (defined as completion and outcomes of TB preventive treatment and TB disease treatment, stratified by individual covariates) held constant at current levels. |
| **Domestic intervention scenarios (international interventions as shown in base-case scenario)** |  |
| Provision of *Mtb* infection testing and treatment for new immigrants (A1) | Provision of *Mtb* infection testing and TB preventive treatment for all new legal immigrants and refugees entering the United States, excluding undocumented migrants or short-term visitors, with 77% acceptance of treatment if indicated (183,000 individuals initiating treatment in first year of scenario). |
| Improved *Mtb* infection testing and treatment within current U.S risk populations (A2) | Increased uptake of *Mtb* infection testing and TB preventive treatment among currently recommended populations in the United States, doubling treatment uptake among previously untreated individuals within each risk group compared with current levels (312,000 additional individuals initiating treatment in first year of scenario), and reducing the fraction discontinuing treatment by 26% through adoption of short-course regimens. |
| Improved TB case detection (A3) | Improved case detection within the United States, with the duration of untreated disease (i.e., time from TB incidence to treatment initiation) reduced by 50% relative to current levels (assumed change from 3.4 months to 1.7 months). |
| Improved TB disease treatment outcomes (A4) | Improved TB treatment effectiveness within the United States, such that treatment discontinuation and failure rates are reduced by 50% from current levels (4.3% to 2.2%). |
| All domestic interventions (A5) | Combination of all domestic service improvements (A1-A4). |
| **International intervention scenarios (domestic interventions as shown in base-case scenario)\*** |  |
| Scale-up of current technology (B1) | Rapid scale-up of current TB interventions within high-burden countries to reach full coverage, including roll-out of modern diagnostics (e.g., Xpert MTB/RIF) and expanded early diagnosis (systematic screening of household contacts and high risk groups), such that by 2030 case detection would identify 95% of individuals developing TB. patient support to maximize treatment completion (e.g., peer support, food supplementation and income generation), scale-up of TB preventive treatment for groups at higher risk of TB disease (100% coverage of contacts of TB cases, individual with HIV, and other key and vulnerable populations from 2022), and expanded access to modern regimens for drug-resistant TB. |
| Scale-up of current and new technology (B2) | As in Scenario B1, with the additional development and introduction of tools enabling the detection and treatment of subclinical TB disease, sufficient to find and treat 20% of individuals with subclinical TB before they develop symptoms. |
| Scale-up of current and new technology, and TB vaccine from 2030 (B3) | As in Scenario B2, with the additional development and introduction of a post-exposure TB vaccine with 60% efficacy, introduced and scaled up in 2030.** |
| **Combined international and domestic intervention scenarios** |  |
| All interventions, excluding new TB vaccine (C1) | *International:* Scale-up of current and new technology internationally, excluding new TB vaccine (B2).<br>*Domestic:* Combination of all domestic service improvements (A5). |
| All interventions, with new TB vaccine internationally (C2) | *International:* Scale-up of current and new technology internationally, including a new TB vaccine (B3).<br>*Domestic:* Combination of all domestic service improvements (A5). |

### Study outcomes

The primary study outcome was TB incidence rates in the overall U.S. population, calculated as the number of new TB cases estimated by the simulation model divided by resident population in that year, expressed per 100,000. We also calculated incidence for U.S.-born and non-U.S.-born residents separately and reported the year in which U.S. TB incidence rates were projected to reach pre-elimination and elimination thresholds.^16^ We calculated additional outcomes quantifying health and economic benefits within the United States, each comparing a given intervention scenario to the base-case: (i) *Mtb* infections averted, (ii) TB deaths averted, (iii) life-years gained, (iv) quality-adjusted life years (QALYs) gained, and (v) the economic value of TB incidence reductions. To calculate QALYs gained, we compared the number of life-years lived in each intervention scenario to the base-case, adjusting for the lower quality of life associated with TB disease.^38^ The economic value of TB incidence reductions was calculated as the sum of averted TB treatment costs, averted productivity losses from TB disease, and the societal value of averted TB mortality estimated via a ‘Value of Statistical Life’ (VSL) approach^39^ (details in Supplement). The primary results were estimated for the 2025-2050 period, given the increased uncertainty of longer-term projections.

### Statistical analysis

We used Monte Carlo simulation to propagate uncertainty in analytic inputs through the analysis.^40^ To do so, we re-estimated results for each of 1,000 parameter sets generated by the Bayesian calibration approach and used a latin hypercube sample to generate values for parameters not included in the calibration (VSL inputs, utility weights, future migration trends). This generated 1,000 values for each outcome. We calculated point estimates for each outcome as the median of these values and calculated 95% uncertainty intervals as the 2.5^th^ and 97.5^th^ percentiles. We calculated partial rank correlation coefficients (PRCCs) describing parameters with the greatest influence over study outcomes, and also re-estimated outcomes for high and low values for future migration rates (+/- 50% compared to the main analysis). Analyses were implemented in R (v4.2.2) and C++ via the Rcpp package (v1.0.9).^41,42^

### Role of the funding source

Employees of the funder participated as study coinvestigators.

## Results

### Base-case epidemiological projections

Figure 1 shows base-case projections of future TB incidence rates within the United States, for U.S.-born and non-U.S.-born persons as well as the overall population. While incidence rates in the U.S.-born population are already below the pre-elimination threshold,^13^ incidence rates in the overall U.S. population were not projected to cross this threshold before 2100. By 2050, incidence rates were projected to be 9.5 (95% uncertainty interval: 8.0, 11.5), 0.33 (0.26, 0.39) and 1.8 (1.5, 2.1) per 100,000 in non-U.S.-born, U.S.-born, and total United States populations, respectively. Annual percentage reductions in incidence rates were higher for U.S.-born compared to non-U.S.-born populations, and these rates of decline flattened over time (Table S2).

**Figure 1:**
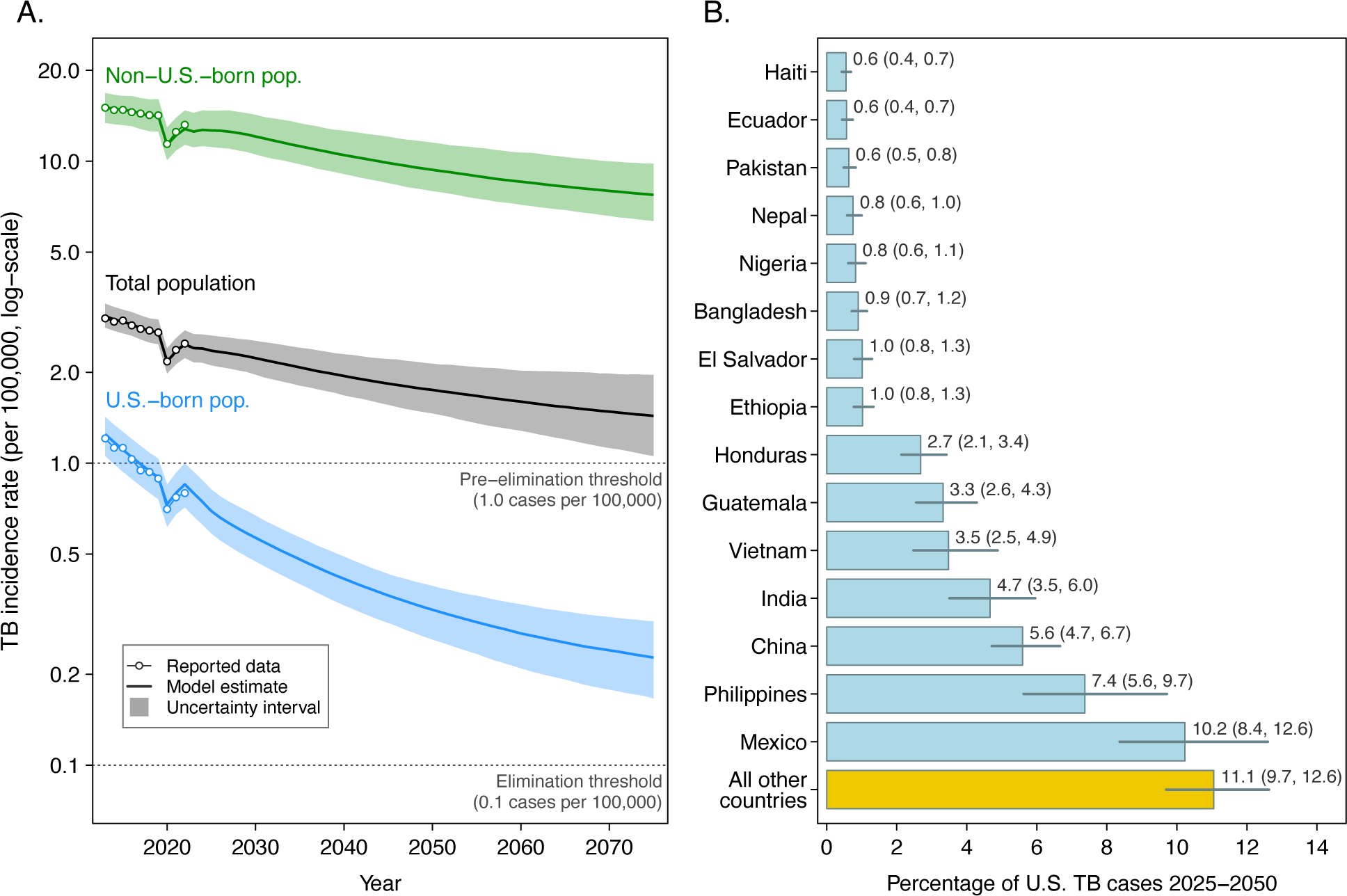
Projections of TB incidence in the United States with continuation of current trends. Panel A: Future TB incidence projections under base-case scenario, with continuation of current TB trends among migrants and TB services coverage and effectiveness within the United States. Panel B: Projected contribution of *Mtb* infection among migrants entering the United States after 2024 as a percentage of total TB cases projected for 2025-2050, by country of birth (top 15 countries shown, plus the combination of all other countries). Values in parentheses represent 95% uncertainty intervals.

TB cases attributable to *Mtb* infections among migrants entering the country after 2024 were projected to make a substantial contribution to future TB incidence. Figure 1 Panel B shows the projected contribution of *Mtb* infections among migrants arriving after 2024 from each birth country to overall U.S. TB cases during 2025-2050. Collectively, these TB cases represented 55% (51, 60) of all TB cases projected for 2025-2050 under the base-case scenario.

### TB incidence trends under intervention scenarios

Each intervention scenario (Table 1) was projected to reduce TB incidence compared to the base-case. Figure 1 Panel A shows projected incidence trends for domestic intervention scenarios. Among these scenarios, interventions focused on treatment of *Mtb* infection (A1, A2) had greater impact on TB incidence rates than those focused on detection and treatment of TB disease (A3, A4). The scenario combining all domestic interventions reduced incidence by 52% (49, 54) in 2050 compared to the base case, and 57% (55, 60) by 2100. For the international scenarios, scaling up new and anticipated TB interventions including a vaccine (B3) globally available in 2030 was projected to reduce TB incidence in the United States by 38% (33, 44) in 2050 compared to the base-case, and 76% (72, 80) by 2100.

**Figure 2:**
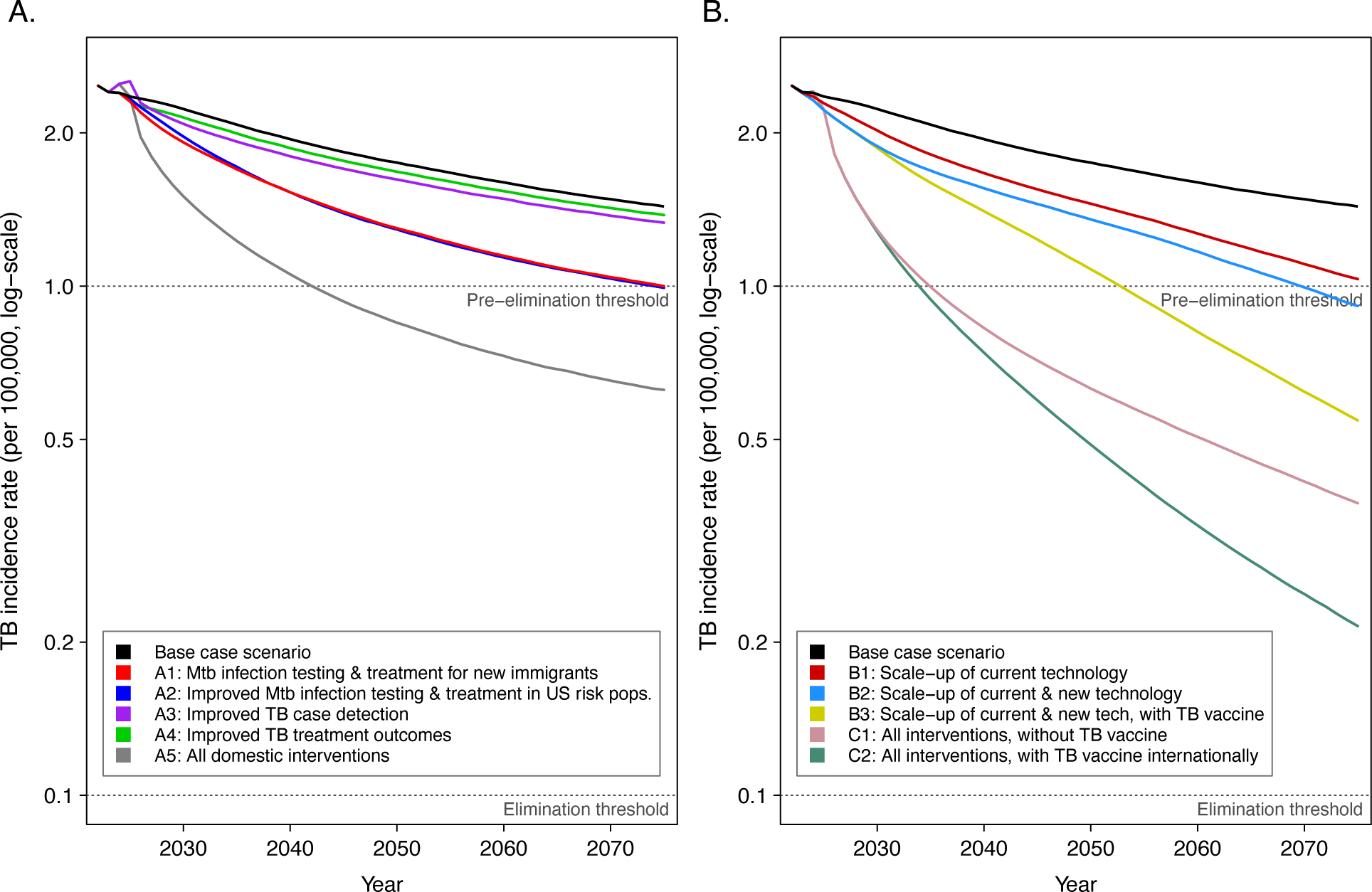
Projections of TB incidence in the United States under domestic and international intervention scenarios. Panel A: Future TB incidence projections under domestic intervention scenarios. Panel B: Future TB incidence projections under international and combined intervention scenarios. Uncertainty intervals omitted for clarity. *Mtb* = *Mycobacterium tuberculosis*.

The combined scenarios were projected to produce the greatest incidence reductions. The combination of all domestic and international interventions (C2) produced an estimated U.S. incidence rate of 0.49 (0.40, 0.63) per 100,000 in 2050, 72% (69, 75) less than under the base-case in the same year, and 80% (75, 83) below the rate estimated for 2024. Combining all interventions except an international TB vaccine (scenario C1), produced an incidence rate of 0.63 (0.51, 0.79) per 100,000 in 2050. With or without a vaccine, the combination of domestic and international scenarios reduced incidence in the United States by >50% by 2035, as compared to the base-case. Table S3 reports incidence rates in selected future years for each intervention scenario.

### Health benefits of intervention scenarios

Each scenario was projected to produce notable improvements in health outcomes compared to the base-case (Table 2). Among the domestic scenarios, those focused on treating *Mtb* infection had the greatest impact on TB incidence, while improved TB case detection (A3) had the greatest impact on other outcomes. Over 2025-2050, the combination of all domestic and international interventions (scenario C2) was projected to avert 351 (292, 433) thousand *Mtb* infections, 101 (84, 120) thousand TB cases, and 13 (11, 16) thousand TB deaths inside the United States. Combining the survival gains resulting from these averted deaths with reductions in morbidity through averted TB cases, this would save 259 (205, 322) thousand QALYs. Overall, the economic value of TB incidence reductions was estimated to be US$68 (33, 112) billion. Table S4 reports these impacts as percentage reductions compared to the base-case. Table S5 reports disaggregated economic outcomes.

**Table 2:** Impact of base-case and intervention scenarios on cumulative health and economic outcomes, 2024-2050. QALYs = quality-adjusted life years, *Mtb* = *Mycobacterium tuberculosis*, USD = U.S. dollars.

|  | *Mtb*<br>infections<br>averted<br>(000s) | TB cases<br>averted<br>(000s) | TB deaths<br>averted<br>(000s) | Life-years<br>saved (000s) | QALYs saved<br>(000s) | Value of TB<br>incidence<br>reductions<br>(USD bil.) |
| --- | --- | --- | --- | --- | --- | --- |
| **Domestic intervention scenarios** |  |  |  |  |  |  |
| *Mtb* infection testing and treatment for new immigrants (A1) | 81<br>(65, 104) | 34<br>(29, 42) | 3.5<br>(2.6, 4.5) | 66<br>(50, 85) | 71<br>(54, 91) | 18<br>(9, 30) |
| Improved *Mtb* infection testing and treatment in U.S. risk populations (A2) | 79<br>(63, 98) | 33<br>(27, 40) | 4.2<br>(3.1, 5.1) | 67<br>(50, 85) | 73<br>(54, 91) | 19<br>(9, 31) |
| Improved TB case detection (A3) | 221<br>(183, 274) | 11<br>(8, 15) | 5.1<br>(4.2, 6.2) | 94<br>(73, 115) | 102<br>(81, 125) | 26<br>(12, 43) |
| Improved TB disease treatment outcomes (A4) | 17<br>(14, 23) | 7<br>(6, 9) | 1.5<br>(1.2, 1.8) | 29<br>(23, 36) | 30<br>(24, 37) | 8<br>(4, 13) |
| All domestic interventions (A5) | 301<br>(249, 373) | 74<br>(63, 87) | 11.5<br>(9.1, 13.9) | 203<br>(159, 250) | 219<br>(174, 269) | 58<br>(28, 93) |
| **International intervention scenarios** |  |  |  |  |  |  |
| Scale-up of current technology (B1) | 65<br>(47, 91) | 23<br>(17, 31) | 2.1<br>(1.4, 3.1) | 42<br>(28, 61) | 46<br>(31, 66) | 12<br>(5, 20) |
| Scale-up of current and new technology (B2) | 93<br>(69, 130) | 34<br>(25, 45) | 3.0<br>(2.0, 4.6) | 62<br>(42, 89) | 67<br>(46, 96) | 17<br>(8, 30) |
| Scale-up of current and new technology, and TB vaccine from 2030 (B3) | 119<br>(92, 161) | 45<br>(36, 59) | 4.1<br>(2.9, 5.8) | 84<br>(60, 115) | 91<br>(66, 124) | 23<br>(11, 39) |
| **Combined international and domestic intervention scenarios** |  |  |  |  |  |  |
| All interventions, excluding new TB vaccine (C1) | 343<br>(285, 422) | 94<br>(78, 112) | 12.9<br>(10.2, 15.8) | 231<br>(181, 288) | 249<br>(197, 309) | 66<br>(32, 108) |
| All interventions, with new TB vaccine internationally (C2) | 351<br>(292, 433) | 101<br>(84, 120) | 13.3<br>(10.5, 16.3) | 240<br>(188, 300) | 259<br>(205, 322) | 68<br>(33, 112) |

### Dates of reaching pre-elimination and elimination thresholds

Under the base-case TB incidence in the United States was not projected to reach the pre-elimination threshold before 2100. Figure 3 shows projected dates for achieving pre-elimination under the combined scenarios. Under each of these scenarios pre-elimination was achieved during the study period. For the most aggressive scenario (C2)—combining all domestic and international interventions—pre-elimination was achieved in 2033 (2031, 2037). The addition of TB vaccination in the international scenarios shortened the time to pre-elimination. Even with the most aggressive scenarios there was less than 50% probability that the elimination threshold of incidence <0.1 per 100,000 would be achieved by 2100 (Table S6).

**Figure 3:**
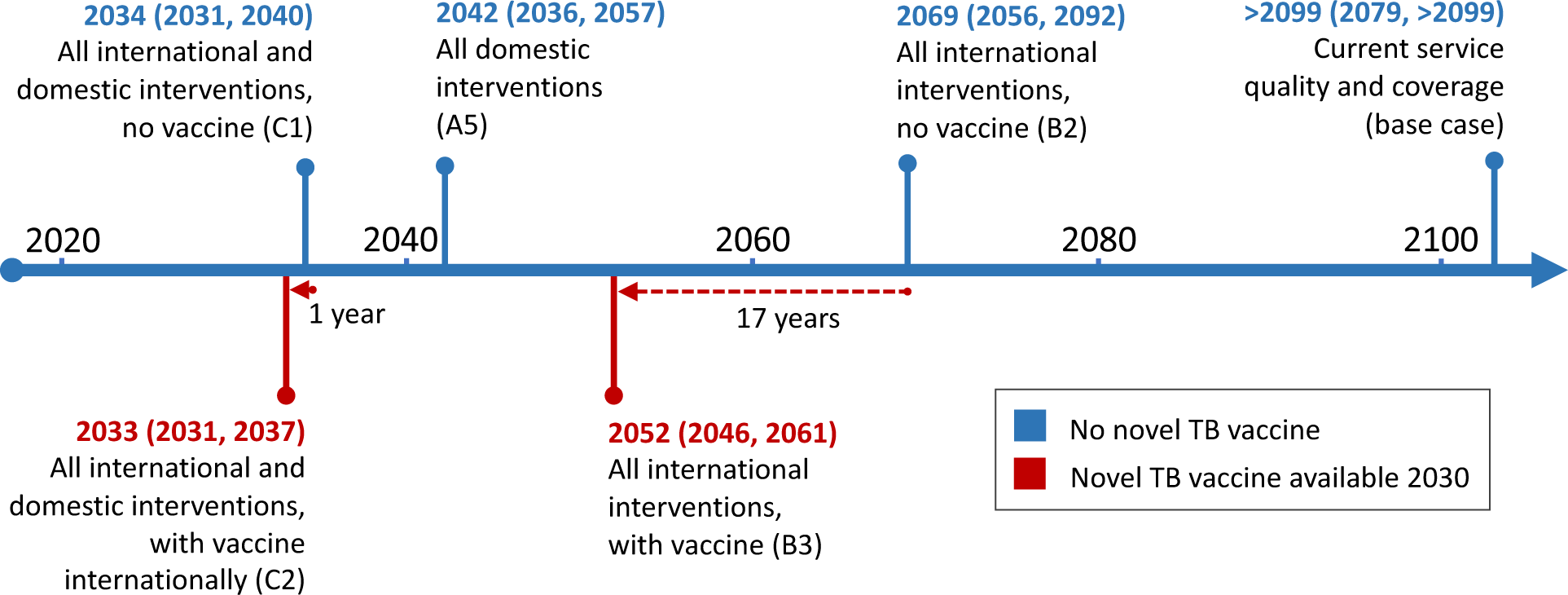
Predicted date of reaching pre-elimination threshold (<1.0 per 100,000) under combined US and international scenarios, with and without a new TB vaccine. TB vaccine scenarios represent a TB vaccine introduced internationally in 2030 with 60% effectiveness and 60% immediate uptake in all international settings. International scenarios assume aggressive scale-up of existing and anticipated TB interventions as described in the Stop TB Partnership’s Global Plan to End TB 2023-2030 ^11^, to achieve WHO End TB Strategy targets of a 95% reduction in TB mortality and a 90% reduction in TB incidence between 2015 and 2035 ^3^. Under a sensitivity analysis in which the coverage and effectiveness of domestic interventions was adjusted to match these End TB Strategy targets U.S. TB incidence rates declined rapidly to reach pre-elimination and elimination targets in 2026 (2025, 2026) and 2073 (2062, >2099) respectively.

### Sensitivity analyses

We examined how results varied with changes in rates of future migration into the United States (Table S7). Overall, results were sensitive to rates of future migration, with a 50% reduction in future migration rates reducing estimated incidence in 2050 by 32-40% depending on the scenario, and a 50% increase in migration rates increasing 2050 incidence by 29-37%. Table S8 reports partial rank correlation coefficients (PRCCs) describing the sensitivity of the base-case incidence projections in 2050 to uncertainty in model parameters. In these results, the three most influential parameters were the annual trend in future migration growth post-2024 (PRCC: 0.70 (0.66, 0.75)), the rate of progression from established *Mtb* infection to TB disease (PRCC: 0.56 (0.51, 0.62)), and the rate of *Mtb* infection testing within recommended groups within the United States (PRCC: −0.48, (−0.53, −0.43)). Under a sensitivity analysis in which the coverage and effectiveness of domestic interventions was adjusted to match the aspirational targets of the End TB Strategy as applied in international scenarios, U.S. TB incidence rates declined rapidly, with pre-elimination and elimination targets reached in 2026 (2025, 2026), and 2073 (2062, >2099) respectively.

## Discussion

This study examined the potential evolution of TB epidemiology in the United States, considering several pathways toward TB pre-elimination and elimination in this low incidence setting. Our analyses projected that if current service coverage and effectiveness are maintained, future U.S. TB incidence will continue to decline, but that this rate of decline will slow over time, with pre-elimination not achieved before 2100.

We found that a range of intervention approaches could accelerate progress towards pre-elimination. Domestically, scenarios describing greater domestic *Mtb* infection testing and treatment or pre-entry testing and treatment of prospective immigrants were found to have approximately equivalent impact. Overall, these strategies produced incidence reductions than strategies focused on detection and treatment of TB disease. In part this resulted from modelling assumptions that the duration of untreated disease within the United States is currently short, and treatment completion high, based on U.S. program indicators showing that TB diagnosis and treatment services are performing well.^13^ However, improved TB case detection was estimated to be effective for reducing TB mortality.

Scenarios examining attainment of End TB Strategy targets outside the United States showed major impacts on U.S. TB trends, highlighting the dependence of U.S. TB epidemiology on *Mtb* infection trends among present and future migrants. Even though we assumed that a TB vaccine would only become available in 2030 (5 years later than assumed by the Global Plan), the scenarios including a TB vaccine showed that the development and universal scale-up of TB vaccine outside of the United States would eventually produce major benefits inside the United States. It is possible that these benefits could be achieved through other interventions (for example, through preventive treatment provided at a similar scale as assumed for the vaccine), but in the Global Plan it was assumed this was the only intervention that could be provided with such broad population coverage.

In general, incidence declines under the international scenarios accelerated more slowly than under the domestic scenarios. However, these scenarios produced ongoing declines in incidence rates that were sustained over time, whereas the annual percentage reductions associated with domestic scenarios flattened towards the end of the projection period. These delayed impacts are consistent with the lagged relationship between TB trends in other countries and their implications within the United States. For scenarios that assumed TB burden among new migrants followed their historical trends (i.e., no improvements in TB services outside the United States), the rate of decline in TB incidence flattened progressively in the later years, with further reductions in U.S. TB incidence constrained by the ongoing entry of individuals with *Mtb* infection. These results can be contrasted with modeling of TB elimination in populations with minimal migration, in which elimination can potentially be achieved by aggressive local intervention alone.^43^

The intervention scenarios greatly reduced the expected time until pre-elimination would be reached, with the scenarios for combined domestic interventions and combined international interventions projected to achieve pre-elimination by 2039 and 2052 respectively. Adopting the full range of intervention approaches (both domestic and international) produced the most rapid declines, achieving pre-elimination by 2033. In contrast, we estimated there was a less than 50% probability that TB elimination would be achieved before 2100, even in the most aggressive intervention scenario in the main analysis. In sensitivity analyses assuming that U.S. TB services could be expanded to meet the End TB Strategy targets for percentage reductions in TB incidence and death by 2035, we were able to construct a domestic intervention scenario achieving TB elimination by 2073.

The results of this analysis suggest greater delays until TB elimination is reached compared to earlier modelled analyses of TB the United States.^29,44,45^ This greater pessimism likely results from the flattening of U.S. TB trends observed over recent years, higher migration rate forecasts for future years, and differences in modelling assumptions. This study is one of the first to examine multiple pathways to achieving TB elimination in a low incidence setting based on aspirational international goals, considering the range of intervention approaches that will likely be required. These scenarios show that while pre-elimination could be achieved relatively rapidly, the elimination threshold (requiring a further 10x reduction in incidence rates) will take many years to achieve even under the most aggressive intervention approach.

This analysis has several limitations. First, we took a deliberately narrow perspective in terms of the geographic scope considered, focusing on outcomes for the U.S. population, even though the intervention strategies included service improvements in other countries. Second, this analysis did not consider the resources required to implement each intervention approach. These costs are expected to be considerable.^11^ Third, this analysis focused on population-level outcomes, and did not consider the individual-level risks and benefits for intervention recipients. It is possible that as TB incidence declines, intervention processes and targeting approaches will need to evolve so that the benefits of an intervention still outweigh the costs and risks for intervention recipients.^46^ Fourth, several scenarios were based on interventions included in the Global Plan to End TB.^11^ These scenarios represent the agreement of global scientific and funding agencies focused on combatting TB and—apart from assuming a slower timeline for vaccine availability—we did not re-estimate the timing or magnitude of these changes. Fifth, the relatively long span of the projections means that future technology development could radically change intervention options.^47^ While our analyses anticipated interventions that could become available in the near term (most notably a new TB vaccine), this required us to make unverifiable assumptions about intervention effectiveness and uptake.^48^ Longer-term technological advances are difficult to anticipate, and may allow more rapid achievement of TB elimination. Finally, this analysis focused on health services improvements, despite evidence that socio-economic factors are also influential determinants of TB trends.^49^ Reducing economic barriers to care, improving housing and living conditions, and addressing under-nutrition all provide further avenues for reaching TB goals, in addition to the strategies examined in this analysis.^50,51^

Achieving TB elimination represents a major challenge, even for low incidence settings like the United States. Strategies that combine a range of approaches are likely needed to achieve this goal, involving strengthening of services within the United States, promoting technological advancements, and supporting action against TB in high-burden countries. Implementation of these combined strategies has the potential to achieve substantial health gains within the United States as well as globally.

## Data Availability

Analytic code and most data inputs available upon request to the corresponding author. Disaggregated TB notification data used for model fitting were obtained from the U.S. National TB Surveillance System (NTSS). The NTSS operates under an Assurance of Confidentiality issued by the Centers for Disease Control and Prevention under Sections 306 and 308(d) of the Public Health Service Act (42 USC 242k and 242m(d)), which prohibits disclosure of information that could be used to directly or indirectly identify patients. More information is available in the data rerelease policy from the Division of Tuberculosis Elimination of the Centers for Disease Control and Prevention (https://wonder.cdc.gov/wonder/help/TB/DTBE-Data-ReRelease-Policy-2022-update-2022-05-09.pdf). A limited data set is available at https://wonder.cdc.gov/TB-v2021.html.

## Declaration of interests

The authors declare no conflicts of interest.

## Author contributions

All authors contributed to the design of the study. SMM and GRBA assisted with data access. NAM conducted the analysis and drafted the manuscript. All authors edited the manuscript.

## Acknowledgements

This project was funded by the Centers for Disease Control and Prevention (CDC), National Center for HIV, Viral Hepatitis, STD, and TB Prevention Epidemiologic and Economic Modeling Agreement (NEEMA; #1NU38PS004651). We thank Maryam Haddad, Andrew Hill, Teresa Puente, Stephanie Su, and Cindy Imai for assistance with this analysis.

## Notes

### Competing Interest Statement

The authors have declared no competing interest.

### Funding Statement

This study was funded by the U.S. Centers for Disease Control and Prevention.

### Author Declarations

The Institutional Review Board of Harvard University determined this research to constitute non human subjects research (IRB22-1328).

## References

1. United Nations General Assembly. Resolution adopted by the General Assembly on 10 October 2018: political declaration of the high-level meeting of the General Assembly on the fight against tuberculosis (A/RES/73/3). New York: United Nations, 2018.

2. United Nations General Assembly. Political Declaration of the High-Level Meeting on the Fight Against Tuberculosis : draft resolution / submitted by the President of the General Assembly (A/78/L.4). New York: United Nations, 2023.

3. World Health Organization. The End TB Strategy 2015. Geneva, Switzerland: World Health Organization, 2016.

4. United Nations General Assembly. Transforming our world: the 2030 agenda for sustainable development (A/RES/70/1). Geneva: United Nations General Assembly, 2015.

5. Sahu S, Ditiu L, Lawson L, Ntoumi F, Arakaki D, Zumla A. UN General Assembly tuberculosis targets: are we on track? The Lancet 2020; 395(10228): 928–30.

6. Dheda K, Perumal T, Moultrie H, et al. The intersecting pandemics of tuberculosis and COVID-19: population-level and patient-level impact, clinical presentation, and corrective interventions. The Lancet Respiratory Medicine 2022; 10(6): 603–22.

7. Boehme CC, Nabeta P, Hillemann D, et al. Rapid molecular detection of tuberculosis and rifampin resistance. New Engl J Med 2010; 363(11): 1005–15.

8. Nyang’wa BT, Berry C, Kazounis E, et al. A 24-Week, All-Oral Regimen for Rifampin-Resistant Tuberculosis. N Engl J Med 2022; 387(25): 2331–43.

9. Sterling TR, Villarino ME, Borisov AS, et al. Three months of rifapentine and isoniazid for latent tuberculosis infection. N Engl J Med 2011; 365(23): 2155–66.

10. Tait DR, Hatherill M, Van Der Meeren O, et al. Final analysis of a trial of M72/AS01E vaccine to prevent tuberculosis. New England Journal of Medicine 2019; 381(25): 2429–39.

11. Stop TB Partnership. The global plan to end TB 2023-2030. Geneva: Stop TB Partnership, 2022.

12. Lönnroth K, Mor Z, Erkens C, et al. Tuberculosis in migrants in low-incidence countries: epidemiology and intervention entry points. The International Journal of Tuberculosis and Lung Disease 2017; 21(6): 624–36.

13. U.S. Centers for Disease Control and Prevention. Reported Tuberculosis in the United States, 2021. Atlanta, GA, 2022.

14. US Preventive Services Task Force. Screening for Latent Tuberculosis Infection in Adults: US Preventive Services Task Force Recommendation Statement. JAMA 2023; 329(17): 1487–94.

15. Lewinsohn DM, Leonard MK, LoBue PA, et al. Official American Thoracic Society/Infectious Diseases Society of America/Centers for Disease Control and Prevention clinical practice guidelines: diagnosis of tuberculosis in adults and children. Clin Infect Dis 2017; 64(2): e1–e33.

16. Lönnroth K, Migliori GB, Abubakar I, et al. Towards tuberculosis elimination: an action framework for low-incidence countries. European Respiratory Journal 2015; 45(4): 928–52.

17. U.S. Centers for Disease Control Prevention. Division of TB Elimination: Strategic Plan 2022–2026. Atlanta GA: U.S. Centers for Disease Control Prevention, 2023.

18. Blomquist ET. Program Aimed at Eradication of Tuberculosis. Public Health Rep 1963; 78: 897–905.

19. Dowdle WR. A strategic plan for the elimination of tuberculosis in the United States. MMWR 1989; 38(3): 1–25.

20. Schildknecht KR, Pratt RH, Feng P-JI, Price SF, Self JL. Tuberculosis—United States, 2022. Morbidity and Mortality Weekly Report 2023; 72(12): 297.

21. Clancy L, Rieder H, Enarson D, Spinaci S. Tuberculosis elimination in the countries of Europe and other industrialized countries. European Respiratory Journal 1991; 4(10): 1288–95.

22. Menzies NA, Hill AN, Cohen T, Salomon JA. The impact of migration on tuberculosis in the United States. Int J Tuberc Lung Dis 2018; 22(12): 1392–403.

23. Vespa J, Armstrong DM, Medina L. Demographic turning points for the United States: Population projections for 2020 to 2060. Washington DC, USA: US Census Bureau, 2018.

24. Menzies NA, Bellerose M, Testa C, et al. Impact of Effective Global Tuberculosis Control on Health and Economic Outcomes in the United States. Am J Respir Crit Care Med 2020; 202(11): 1567–75.

25. Schwartzman K, Oxlade O, Barr RG, et al. Domestic Returns from Investment in the Control of Tuberculosis in Other Countries. New Engl J Med 2005; 353(10): 1008–20.

26. Menzies NA, Shrestha S, Parriott A, et al. The Health and Economic Benefits of Tests That Predict Future Progression to Tuberculosis Disease. Epidemiology 2022; 33(1): 75–83.

27. Menzies NA, Parriott A, Shrestha S, et al. Comparative Modeling of Tuberculosis Epidemiology and Policy Outcomes in California. Am J Respir Crit Care Med 2020; 201(3): 356–65.

28. Shrestha S, Parriott A, Menzies NA, et al. Estimated Population-Level Impact of Using a Six-Week Regimen of Daily Rifapentine to Treat Latent Tuberculosis Infection in the United States. Ann Am Thorac Soc 2020; 17(12): 1639–42.

29. Menzies NA, Cohen T, Hill AN, et al. Prospects for Tuberculosis Elimination in the United States: Results of a Transmission Dynamic Model. Am J Epidemiol 2018; 187(9): 2011–20.

30. Ruggles S, Flood S, Sobek M, et al. IPUMS USA: Version 13.0 [10.18128/D010.V13.0] Minneapolis MN, USA: University of Minnesota, 2023.

31. Menzies NA. High-resolution estimates of the foreign-born population and international migration for the United States. arXiv preprint arXiv:190601716 2019.

32. Menzies NA, Soeteman DI, Pandya A, Kim JJ. Bayesian methods for calibrating health policy models: a tutorial. Pharmacoeconomics 2016; 35(6): 613–24.

33. Raftery AE, Bao L. Estimating and projecting trends in HIV/AIDS generalized epidemics using Incremental Mixture Importance Sampling. Biometrics 2010; 66(4): 1162–73.

34. WHO Global TB Programme. WHO Global TB Database [http://www.who.int/tb/country/data/download/en/]. Geveva Switzerland: WHO Global TB Programme; 2023.

35. Houben RM, Dodd PJ. The Global Burden of Latent Tuberculosis Infection: A Re-estimation Using Mathematical Modelling. PLoS Med 2016; 13(10): e1002152. doi: 10.1371/journal.pmed. eCollection 2016 Oct.

36. Miramontes R, Hill AN, Yelk Woodruff RS, et al. Tuberculosis Infection in the United States: Prevalence Estimates from the National Health and Nutrition Examination Survey, 2011-2012. PLOS ONE 2015; 10(11): e0140881.

37. U.S. Centers for Disease Control and Prevention. Online Tuberculosis Information System (OTIS), National Tuberculosis Surveillance System, United States, 1993-2021 [https://wonder.cdc.gov/TB-v2021.html]. Atlanta GA, USA: U.S. Centers for Disease Control and Prevention, 2023.

38. Bauer M, Ahmed S, Benedetti A, et al. The impact of tuberculosis on health utility: a longitudinal cohort study. Qual Life Res 2015; 24(6): 1337–49.

39. Robinson LA HJ, Baxter JR. Guidelines for regulatory impact analysis [https://aspe.hhs.gov/pdf-report/guidelines-regulatory-impact-analysis]. Washington DC: US Department of Health and Human Services, 2016.

40. Briggs AH, Weinstein MC, Fenwick EAL, Karnon J, Sculpher MJ, Paltiel AD. Model parameter estimation and uncertainty: a report of the ISPOR-SMDM Modeling Good Research Practices Task Force--6. Value Health 2012; 15(6): 835–42.

41. Eddelbuettel D, Francois R. Rcpp: Seamless R and C++ Integration. J Stat Softw 2011; 40(8): 1–18.

42. R Core Team. R: A language and environment for statistical computing [https://www.R-project.org]. Vienna, Austria: R Foundation for Statistical Computing, 2022.

43. Hill P, Dye C, Viney K, et al. Mass treatment to eliminate tuberculosis from an island population. The International journal of tuberculosis and lung disease 2014; 18(8): 899–904.

44. Dye C, Glaziou P, Floyd K, Raviglione M. Prospects for tuberculosis elimination. Annual review of public health 2013; 34.

45. Hill AN, Becerra JE, Castro KG. Modelling tuberculosis trends in the USA. Epidemiol Infect 2012; 140: 1862–72.

46. Schluger NW. Tuberculosis elimination, research, and respect for persons. American Thoracic Society; 2019. p. 560–3.

47. Salomon JA, Weinstein MC, Goldie SJ. Taking account of future technology in cost effectiveness analysis. BMJ 2004; 329(7468): 733–6.

48. Nelson KN, Shah NS, Cranmer LM, Vasudevan L, Bednarczyk RA. An effective vaccine is only the first step: the need to create and sustain demand for TB vaccines. Int J Tuberc Lung Dis 2023; 27(10): 718–20.

49. Lönnroth K, Jaramillo E, Williams BG, Dye C, Raviglione M. Drivers of tuberculosis epidemics: the role of risk factors and social determinants. Soc Sci Med 2009; 68(12): 2240–6.

50. Carter DJ, Glaziou P, Lönnroth K, et al. The impact of social protection and poverty elimination on global tuberculosis incidence: a statistical modelling analysis of Sustainable Development Goal 1. The Lancet Global Health 2018; 6(5): e514–e22.

51. Bhargava A, Bhargava M, Meher A, et al. Nutritional supplementation to prevent tuberculosis incidence in household contacts of patients with pulmonary tuberculosis in India (RATIONS): a field-based, open-label, cluster-randomised, controlled trial. Lancet 2023; 402(10402): 627–40.

