## Supplementary materials for "The long-term impacts of domestic and international TB service improvements on TB trends within the United States: a mathematical modelling study"

Supplementary methods: details on specification of analytic scenarios

***Domestic scenarios***: Domestic base-case and intervention scenarios were based on scenarios examined in a prior modelling study (1). We simulated coverage and effectiveness of TB interventions to match current data. For *Mtb* infection, this included past scale-up of targeted testing and treatment within recommended risk groups to match published coverage estimates (2). In the United States, targeted testing and treatment for *Mtb* infection is recommended for individuals with elevated risks of developing TB, either due greater exposure to infection (birth or former residence in countries with high TB prevalence, current or former residence in high-risk congregate settings including homeless shelters and correctional facilities, and contact with individuals with infectious TB disease), or elevated TB progression rates (evidence of recent *Mtb* infection or immunosuppressive conditions/medications (HIV, organ transplant recipients, other immunosuppressive medications)) (3, 4). Diagnosis was assumed to be via Interferon-Gamma Release Assay (IGRA), with sensitivity and specificity values taken from a latent class analysis of data from the U.S. TB Epidemiologic Studies Consortium (5). We assumed 77% acceptance of treatment among individuals testing positive for *Mtb* infection, based on national data on uptake of treatment for *Mtb* infection among close contacts of infectious TB cases (6). Under the base-case scenario we assumed that uptake of *Mtb* infection screening and treatment resulted in 333,000 (95% uncertainty interval: 255,000, 402,000) individuals initiated a treatment regimen in 2024. *Mtb* infection treatment was assumed to be provided with one of several drug regimens (a 4-month regimen of rifampin (4R, 60%) (7), a 12-week regimen of isoniazid and rifapentine (3HP, 21%) (8), and a 6-month regimen of isoniazid (6H, 19%) (9)), reflecting recent U.S. prescribing patterns (10). Treatment completion was assumed to be 87% for 3HP (11) and lower for other regimens, assuming a constant monthly rate of discontinuation over the length of each regimen (average completion 83%). Treatment efficacy was assumed to be 93% for individuals completing each regimen (7-9), and 0% otherwise. Under intervention scenario A1 (*provision of Mtb infection testing and treatment for new immigrants*) we assumed screening and treatment would be provided before entry the United States by U.S. panel physicians (12), and would not apply to short-term visitors and undocumented migrants. Treatment acceptance and completion was assumed to be 67% and 88% respectively, based on a pilot study of *Mtb* infection testing and treatment offered to US immigrant visa applicants in Vietnam (13). In the first year of the intervention scenario 183,000 (152,000, 218,000) individuals were simulated as initiating treatment via this intervention. We assumed treatment would be with 3HP, and other features of testing and treatment are as described above for the base-case scenario.

Under intervention scenario A2, *(improved Mtb infection testing and treatment within current U.S risk populations*) we assumed a doubling of the rate of *Mtb* infection screening among recommended risk groups described under the base-case scenario, with an additional 312,000 (240,000, 375,000) individuals initiating treatment in the first year of the intervention scenario (2025) compared to the base-case (79% non-U.S.-born, 2% with HIV, 1% homeless, and 18% from contacts of individuals with infectious TB and other risk populations). For this scenario we also assumed that all individuals would receive 3HP for *Mtb* infection treatment, reducing treatment non-completion by 26% (from 17% to 13%). For individuals developing TB disease, the base-case scenario assumed diagnosis and treatment would match published treatment outcomes data (14). Intervention scenario A3 (*improved TB case detection*) assumed that the rate at which individuals with untreated TB disease would present for healthcare, receive a correct diagnosis and be initiated on treatment would be doubled, effectively reducing the duration of untreated disease by 50% relative to current levels (1.7 months compared with 3.4 months (fitted value from model calibration)), shortening the infectious period and reducing pre-treatment morbidity and mortality. Intervention scenario A4 (*improved TB disease treatment outcomes*) assumed improvements in the clinical care of diagnosed TB disease, such that treatment discontinuation and failure rates would be 50% lower than current levels (from 4.3% to 2.2%) (14). Apart from intervention scenario A1, all activities associated with domestic intervention scenarios were assumed to occur within the United States.

***International scenarios***: International base-case and intervention scenarios were based on scenarios examined in the Stop TB Partnership's Global Plan to End TB 2023-2030 (15). In modelling conducted for the Global Plan, cubic spline regression was used to project baseline trends for each country through recent incidence and notification data reported by the WHO Global TB Program (16). We used this baseline projection as our base-case scenario. The full Global Plan scenario involved aggressive scale-up of existing TB interventions within each country to achieve outcomes of outcomes of 95% reduction in TB mortality and 90% reduction in TB incidence rates between 2015 and 2035 (17). These include earlier TB diagnosis via uptake of currently-available rapid molecular diagnostics and systematic screening of household contacts and high-risk groups; improvements in TB cure via universal drug susceptibility testing, better regimens and treatment monitoring as well as patient support via peer support, food supplementation and income generation; improved management of comorbidities; provision of care for post-TB sequalae; scale-up of screening and treatment for *Mtb* infection among child, adolescent and adult contacts of TB cases, individual with HIV, and other key and vulnerable populations depending on epidemiological context; and strengthening of infection prevention and control measures across the health system. The Global Plan also anticipated the introduction of new diagnostic, therapeutic, and vaccine technologies, including a two-dose, post-exposure TB vaccine with 60% efficacy assumed to become available in 2025 and reaching at least 60% of adults and adolescents (>10 years of age) by 2028, and maintaining 60% coverage or more in subsequent years. The mix of interventions, timing of introduction, and extent of scale-up was tailored to each country context. Modelling for the Global Plan was undertaken with a transmission-dynamic TB model fit to country data (18). Additional modelling was conducted to separate out the impact of existing interventions, new technologies apart from a vaccine (modeled as increased detection and treatment of subclinical TB), and the TB vaccine. We used the results of these scenarios to create our three international intervention scenarios (*scale-up of current technology* (B1), *scale-up of current and new technology excluding a TB vaccine* (B2), and *scale-up of current and new technology including a TB vaccine internationally* (B3)), with the adaptation that for our TB vaccine scenario (B3) we assumed that TB vaccine impacts would be delayed until 2030, given the anticipated timeline of vaccine development and roll-out for products in the TB vaccine development pipeline (19). To reproduce Global Plan results we employed an approach used in an earlier study, fitting models to Global Plan epidemiological projections for each individual country and scenario (20). Figure S7 shows incidence projections for each modelled setting under the international intervention scenarios, as well as incidence values in 2015 and 2035 for each setting and scenario. All activities associated with international intervention scenarios were assumed to occur outside of the United States.

<u>Supplementary methods: details on QALY and economic benefit calculations</u>

***Quality-adjusted life years (QALYs)***: this outcome represents the number of QALYs lost in a given year due to TB morbidity and mortality, calculated as the sum of life-years lost to TB, plus reductions in quality of life for individuals with TB disease. Life-years lost to TB were calculated as the total number of TB deaths in each age group multiplied by life expectancy for that age group (21), summed across all age groups. Reductions in quality of life for individuals with TB disease were calculated as the product of TB incidence in each year and the total non-fatal utility loss per TB case. We assumed a mean value of 0.149 (95% interval: 0.075, 0.224) for this input, estimated as a 0.087 utility decrement averaged over the 12 months following treatment initiation, plus a utility decrement of 0.25 applied to the assumed duration of untreated disease prior to treatment initiation (1.7 months for scenarios that assumed expanded domestic TB case detection, 3.4 months otherwise). For simplicity we assumed no utility loss for individuals with *Mtb* infection or receiving TB preventive treatment.

***Economic value of TB incidence reductions***: this outcome represents the total economic value of health gains produced by the intervention scenarios. This outcome was calculated as the sum of (i) averted costs due to reduced TB treatment needs, (ii) reductions in productivity losses from TB disease, and (iii) the societal value of averted TB mortality. Unit costs for TB treatment (including any required hospitalization) were derived from Castro et al. (22) and Winston et al. (23), and updated to 2022 price levels using personal consumption expenditure for health (PCE-Health, inpatient and outpatient indices) and updated medication price lists (24), producing values of $24,512, $172,911, and $599,116 for non-MDR-TB, MDR-(non-XDR)-TB, and XDR-TB respectively. Based on 2021 notification data these regimens were assumed to represent 98.6%, 1.1%, and 0.3% of total cases respectively (14). We applied a 3% discount rate to TB treatment costs incurred in future years (25).

Productivity losses due to TB were calculated as the monetary value of time lost during TB disease treatment and due to TB death. Estimates of annual and lifetime labor productivity for different age groups were derived from Grosse et al. (26). We adjusted these values for inflation using the Consumer Price Index for All Urban Consumers (CPI-U) (27), and adjusted for changes in real income using Current Population Survey (CPS) data on weekly earnings (28). We applied these costs to the modelled estimates of individuals developing TB disease. The percentage of TB patients who are hospitalized (49%) and mean length of stay per patient (24 days) were derived from Taylor et al. (29). Time losses due to outpatient TB treatment were based on Shepardson et al. (30). Annual and lifetime productivity estimates for future years were adjusted for projected real income growth of 0.8% per year (31), and discounted at 3% (25).

The societal value of averted TB mortality was calculated based on standard approaches for Benefit-Cost Analysis adopted by the U.S. Government Department of Health and Human Services (32), whereby the number of averted deaths is multiplied by the value of a statistical life (VSL). An initial VSL value of $9.0 (4.2, 13.7) million in 2013 was used (33), and adjusted to 2022 values accounting for price inflation and changes in real income (34). We adjusted for inflation using the Consumer Price Index for All Urban Consumers (CPI-U) (27). We adjusted for changes in real income using Current Population Survey (CPS) data on weekly earnings (28). This produced a 2022 VSL value of $12.4 (5.8, 18.8) million. For the VSL value for deaths occurring in future years we assumed future real income growth of 0.8% per year (31), and applied a 3% discount rate (25). As TB deaths are concentrated in older age groups with lower remaining life expectancy, using a standard VSL value may over-estimate the welfare loss associated with a TB death. For this reason, we adopted a VSLY (Value of a Statistical Life Year) approach, weighting the value of each death proportional to the discounted remaining life expectancy at the age of death. With these adjustments (accounting for the older age of TB death, future real income growth, and discounting) the average VSL per TB death was US$5.1 (2.3, 7.7) million under the base-case scenario, for the 2025-2050 period.

We summed the averted costs from reduced TB treatment needs, reduced productivity losses from TB disease, and societal value of averted TB deaths to calculate the total economic value of TB incidence reductions. In this calculation we excluded the productivity losses associated with TB mortality to avoid double-counting, as these are implicitly included in the VSL estimates used to calculate the societal value of averted TB deaths.

| Country | N* | Country | N | Country | N |
|---|---|---|---|---|---|
| India | 2604 | Somalia | 491 | Cameroon | 185 |
| Mexico | 2487 | Nigeria | 477 | Afghanistan | 153 |
| Philippines | 2076 | Bhutan | 406 | Indonesia | 153 |
| Viet Nam | 1288 | Bangladesh | 351 | Eritrea | 152 |
| China | 1173 | El Salvador | 348 | Thailand | 152 |
| Guatemala | 928 | Pakistan | 331 | Colombia | 137 |
| Ethiopia | 905 | Kenya | 296 | Sierra Leone | 120 |
| Haiti | 801 | Dominican Republic | 272 | Brazil* | 99 |
| Honduras | 763 | Peru | 255 | High-income countries | 241 |
| Myanmar | 760 | Ecuador | 242 | Other LMIC | 3033 |
| Nepal | 616 | Liberia | 188 | | |

**Table S1: Countries and country groups included in models used to estimate *Mtb* infection prevalence among current and future migrants.** N = number of reported TB cases in the United States for individuals from each country of birth over the 2010-2020 period. LMIC = low and middle-income country.

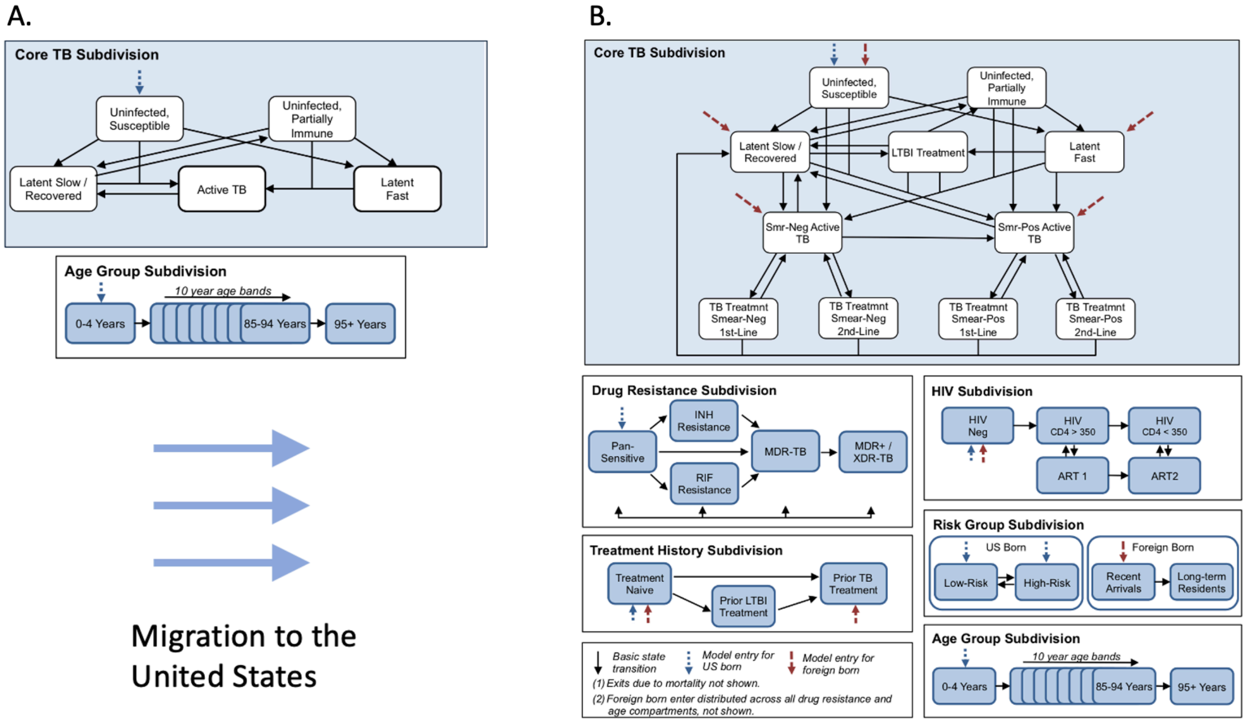

**Figure S1: Schematic of modelled health states and transitions for 32 non-U.S. models (Panel A) and U.S. model (Panel B).** Rounded rectangles indicate modelled health states, arrows represent possible transitions between health states.

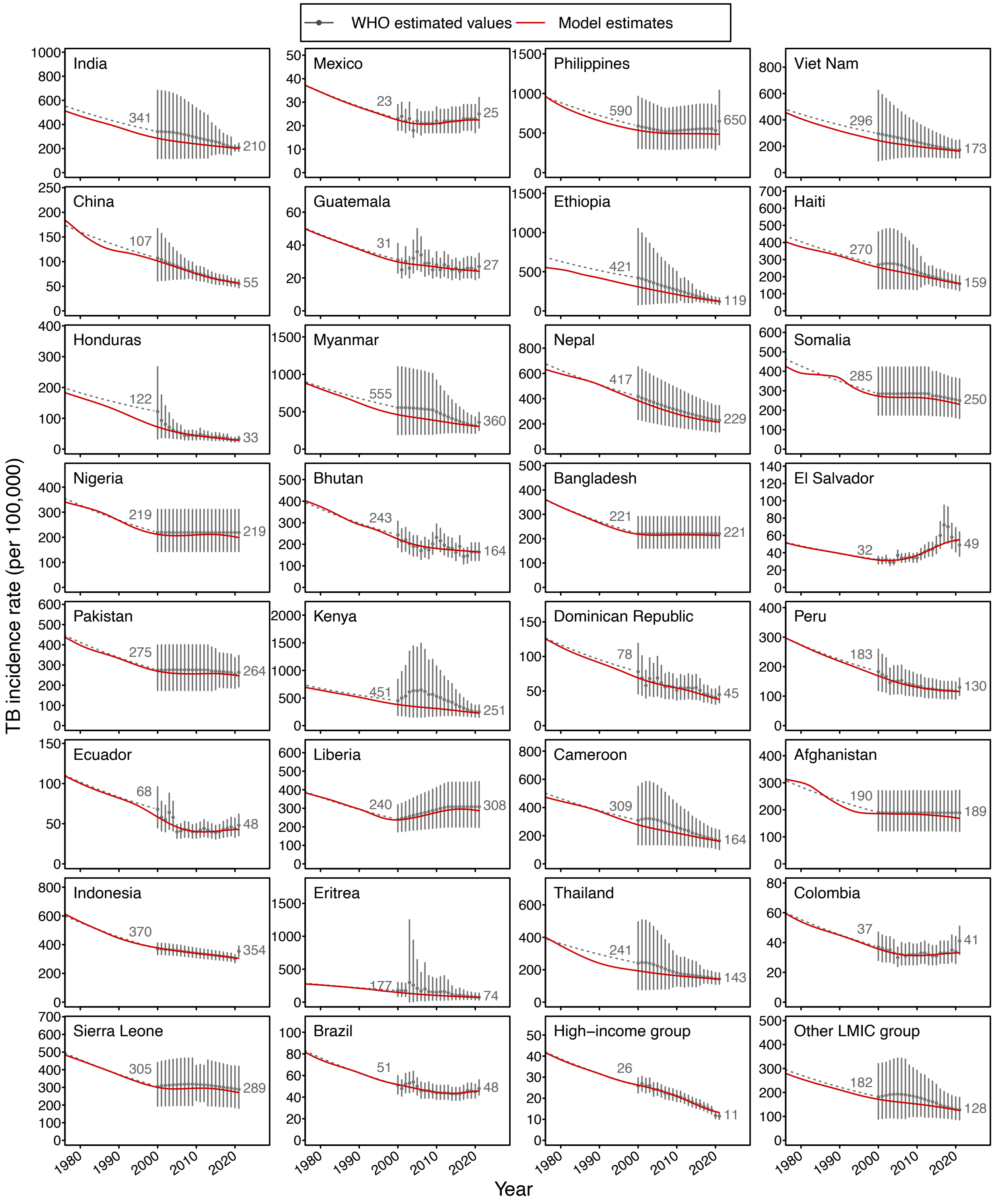

**Figure S2: TB incidence estimates for each non-U.S. country or region of birth over 1980-2021, with calibration of modelled values to WHO estimates**. Figures scaled to range of data. LMIC = low and middle-income country. WHO estimates obtained from WHO Global TB Database (16).

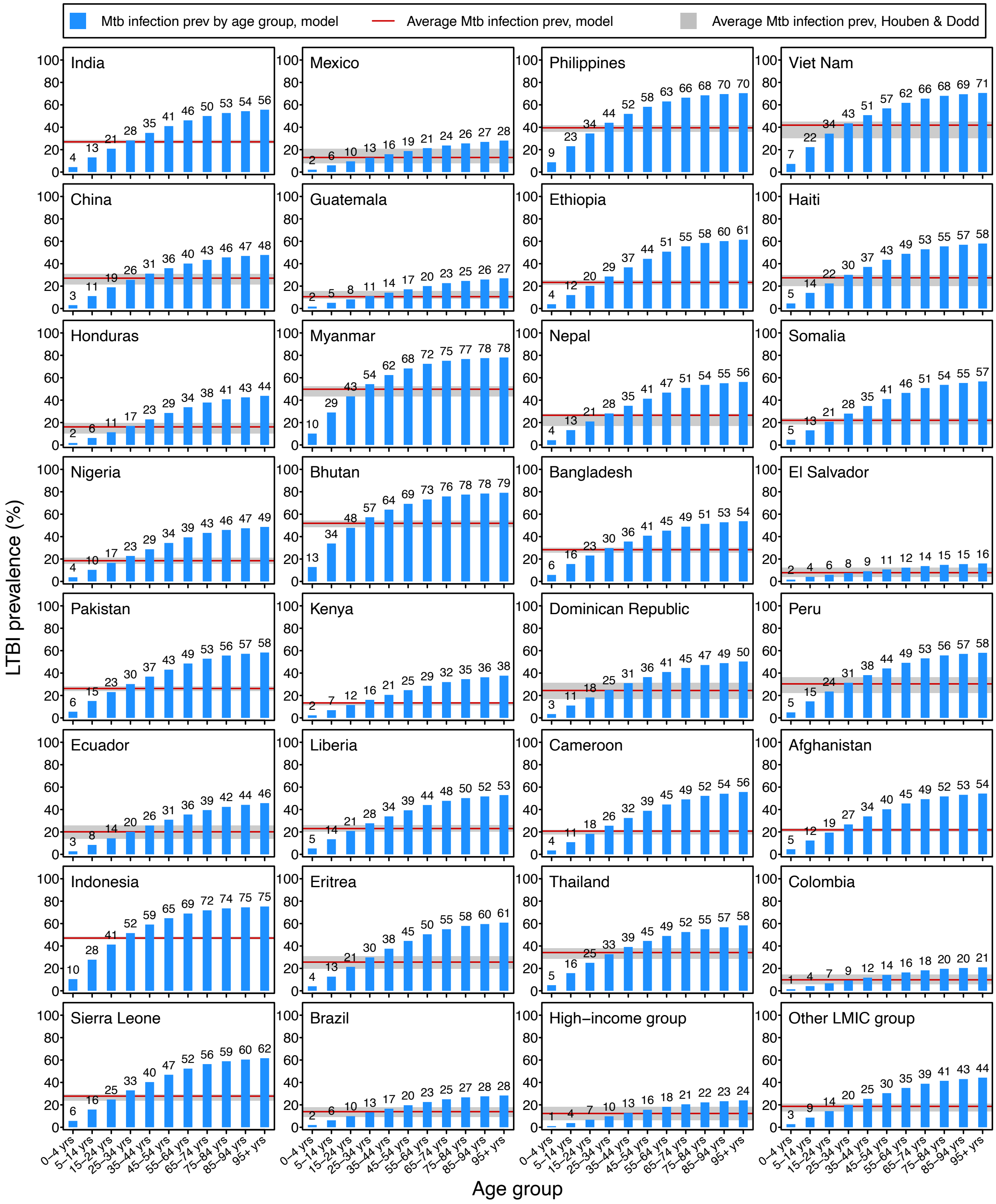

**Figure S3: *Mtb* infection prevalence estimates for each non-U.S. country or region of birth in 2015, calibration of modelled values to published *Mtb* infection prevalence estimates.** LMIC = low and middle-income country. *Published* Mtb infection prevalence estimates drawn from Houben and Dodd 2016 (35).

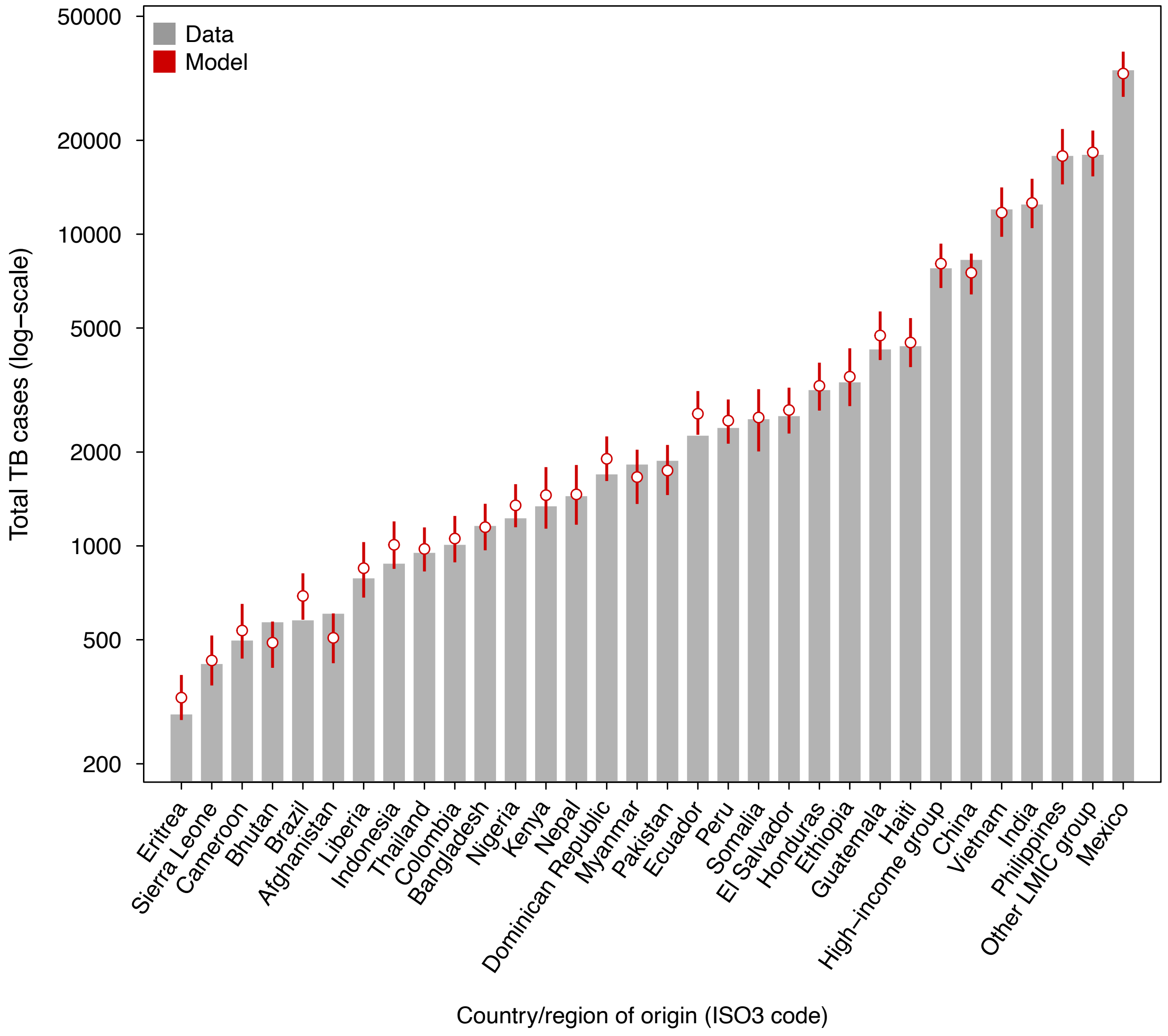

**Figure S4: Number of reported TB cases among non-U.S.-born individuals in the United States between 2000 and 2021 for each country or region of birth, with calibration of modelled values to reported surveillance data.** For modelled values circles represent point estimates, bars represent 95% uncertainty intervals. LMIC = low and middle-income country.

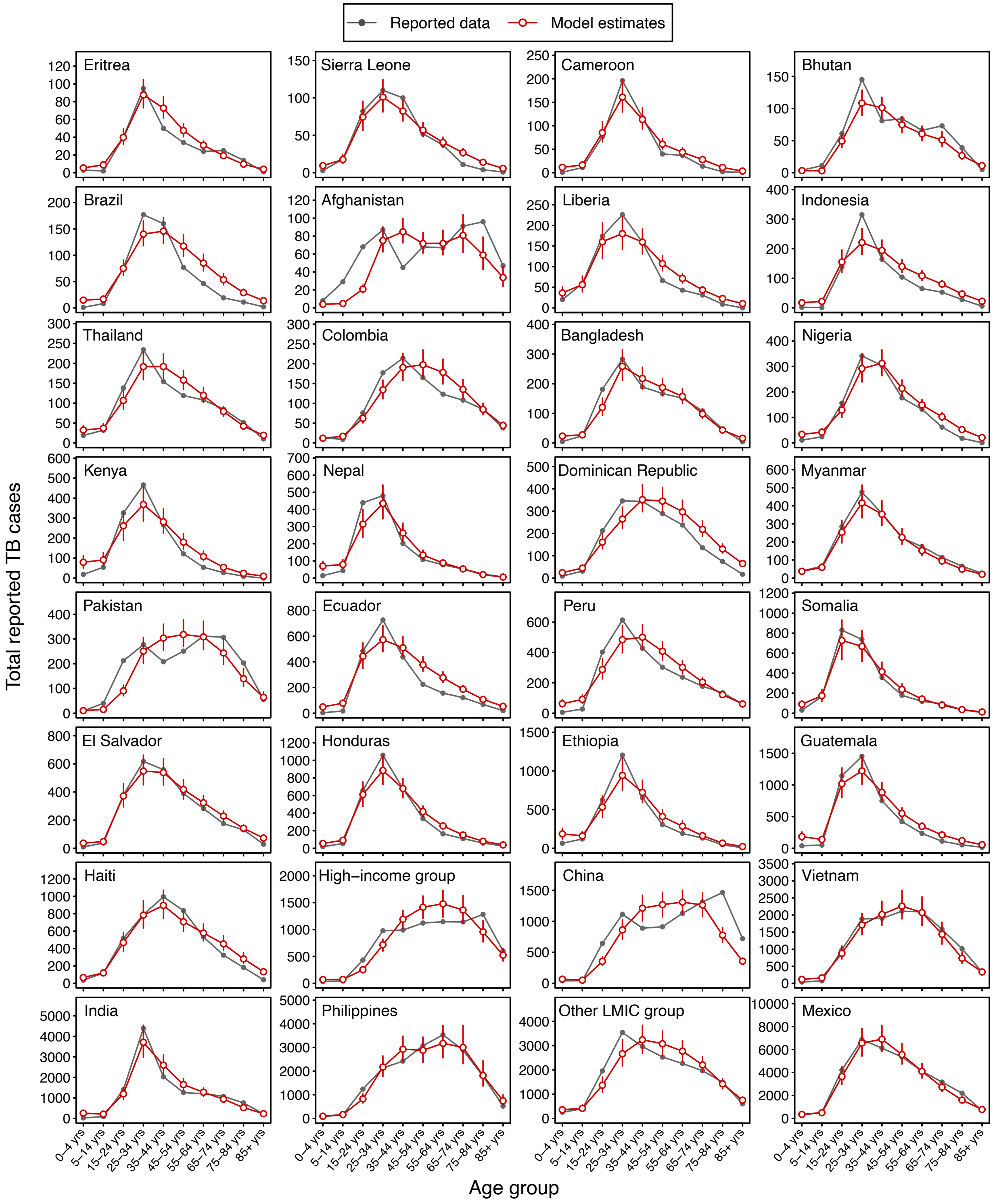

**Figure S5: Number of reported TB cases among non-U.S.-born individuals in the United States between 2000 and 2021 by age group and country/region of birth, with calibration of modelled values to reported surveillance data.** For modelled values circles represent point estimates, bars represent 95% uncertainty intervals. Figures scaled to range of data. LMIC = low and middle-income country.

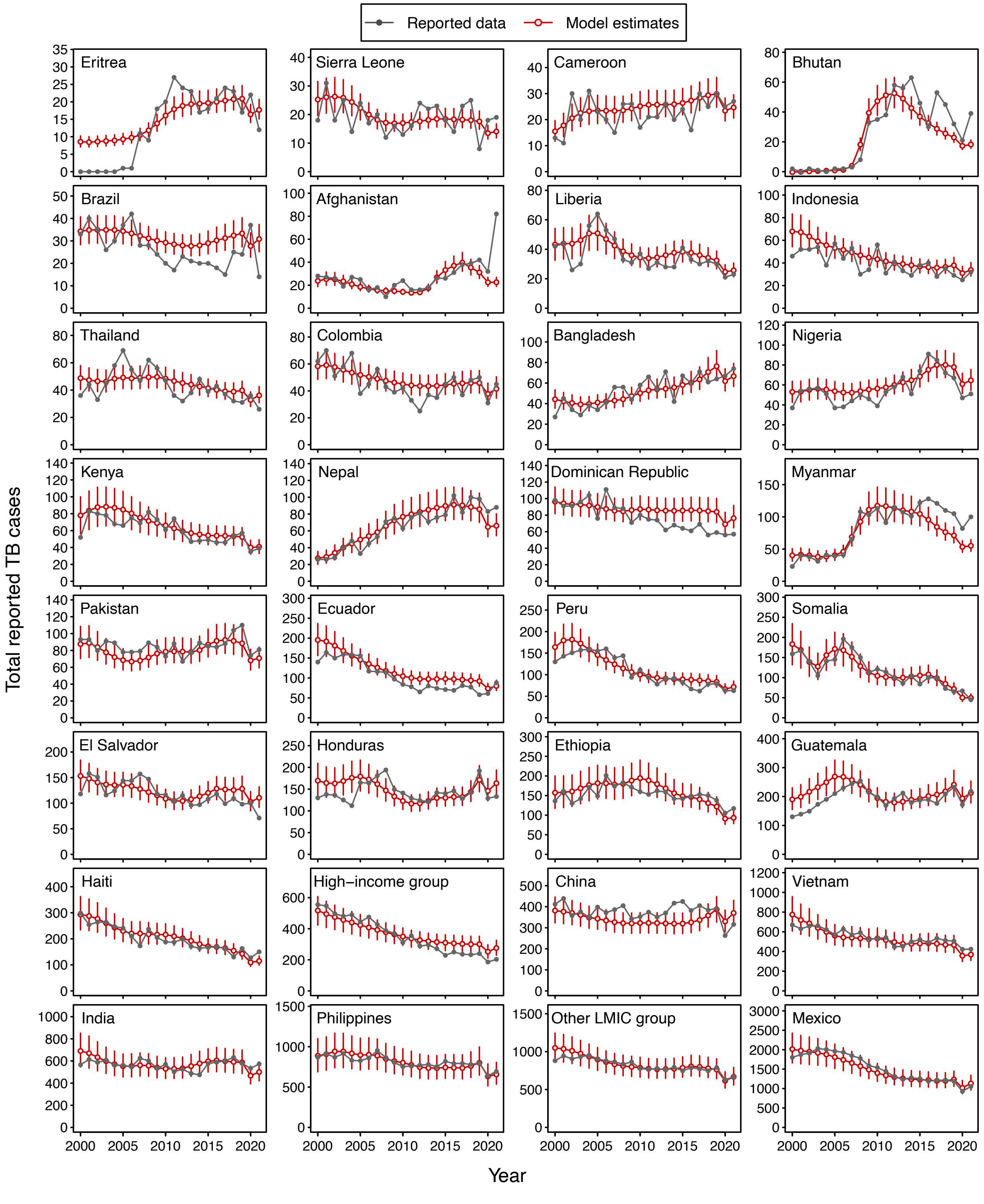

**Figure S6: Number of reported TB cases among non-U.S.-born individuals in the United States between 2000 and 2021 by year and country/region of birth, with calibration of modelled values to reported surveillance data.** For modelled values circles represent point estimates, bars represent 95% uncertainty intervals. Figures scaled to range of data. LMIC = low and middle-income country.

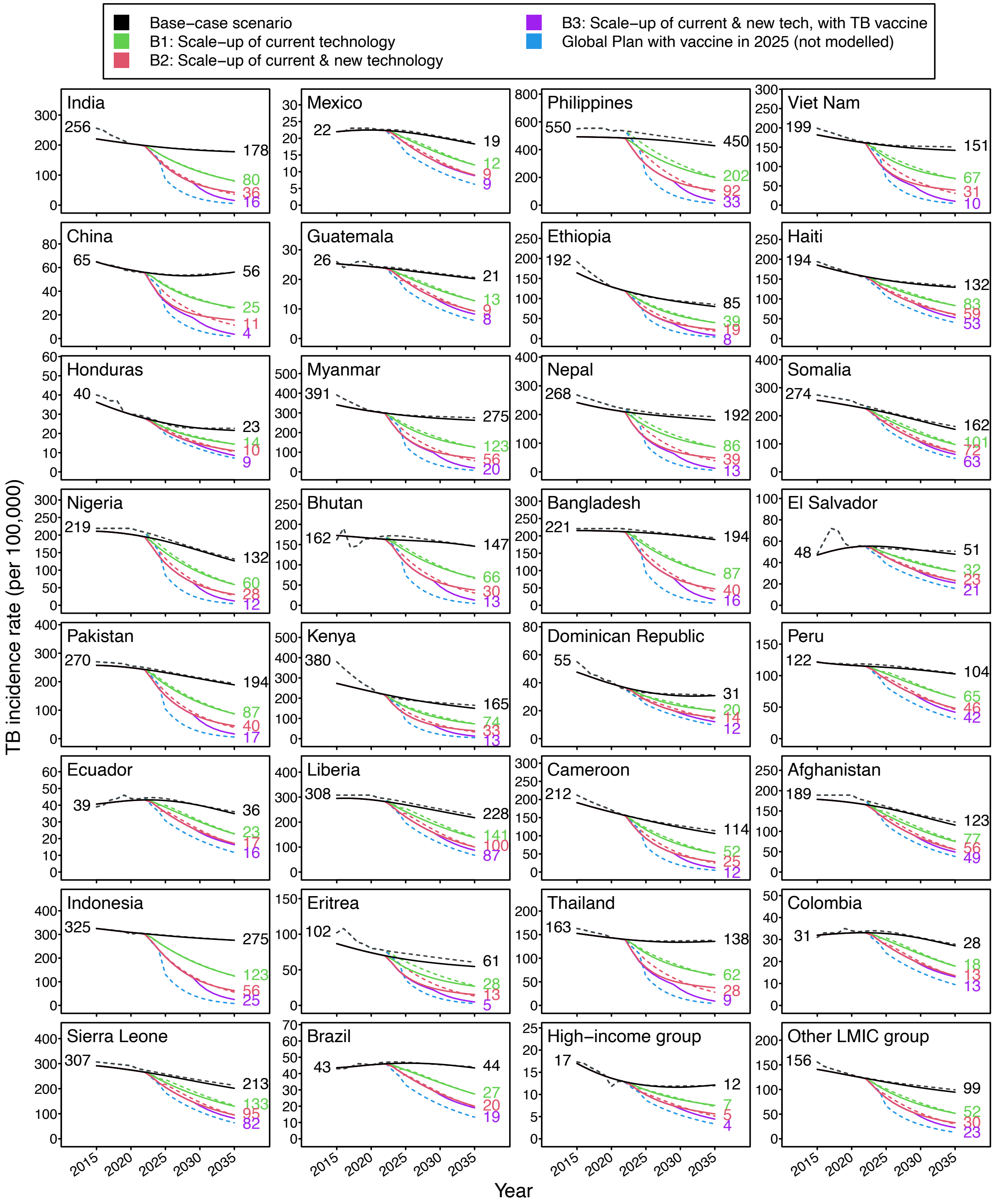

**Figure S7: TB incidence estimates for international intervention scenarios in each non-U.S. country or region of birth over 2015-2035**. Figures scaled to range of data. Scenarios constructed to match projections produced for the Stop TB Partnership's Global Plan to End TB, which is designed to achieve WHO End TB Strategy targets of a 95% reduction in TB deaths and a 90% reduction in TB incidence between 2015 and 2035. Dashed lines represent Global Plan results, solid lines represent model simulations. Scenario A3 based on full Global Plan scenario for each country, except with TB vaccine introduction in 2030 instead of 2025. LMIC = low and middle-income country.

| Year | Non-U.S.-born population | | U.S.-born population | | Total population | |
|---|---|---|---|---|---|---|
| | Incidence rate (per 100,000) | Annual percentage decline (%) | Incidence rate (per 100,000) | Annual percentage decline (%) | Incidence rate (per 100,000) | Annual percentage decline (%) |
| 2035 | 11.33 (9.78, 13.35) | 1.38 (0.39, 2.35) | 0.48 (0.40, 0.57) | 3.04 (2.46, 3.62) | 2.09 (1.81, 2.41) | 1.35 (0.27, 2.27) |
| 2050 | 9.48 (7.96, 11.52) | 1.01 (0.35, 1.63) | 0.33 (0.26, 0.39) | 2.02 (1.36, 2.61) | 1.76 (1.46, 2.13) | 1.00 (0.12, 1.84) |
| 2075 | 7.83 (6.34, 9.82) | 0.54 (0.07, 1.04) | 0.23 (0.17, 0.30) | 0.99 (0.28, 1.72) | 1.45 (1.06, 1.96) | 0.62 (-0.22, 1.38) |
| 2100 | 7.15 (5.60, 9.11) | 0.27 (-0.21, 0.70) | 0.19 (0.12, 0.29) | 0.51 (-0.26, 1.29) | 1.32 (0.80, 1.98) | 0.34 (-0.49, 1.18) |

**Table S2: Projected future incidence rate and annual percentage decline in incidence rate for non-U.S.-born, U.S.-born, and total United States population for selected years.** Values in parentheses represent 95% uncertainty intervals. Annual percentage declines averaged over the +/- 2 years around each selected year.

| | TB incidence rate in given year (per 100,000) | | | | Reduction in incidence rate compared to base-case scenario in same year (%) | | | | Reduction in incidence rate compared to base-case scenario in 2024 (%) | | | |
|---|---|---|---|---|---|---|---|---|---|---|---|---|
| **Scenario** | **2035** | **2050** | **2075** | **2100** | **2035** | **2050** | **2075** | **2100** | **2035** | **2050** | **2075** | **2100** |
| Base-case scenario (continuation of current trends) | 2.08 (1.81, 2.41) | 1.75 (1.46, 2.13) | 1.43 (1.06, 1.96) | 1.28 (0.80, 1.98) | Reference | Reference | Reference | Reference | 13 (6, 19) | 27 (16, 36) | 40 (22, 54) | 46 (19, 65) |
| ***Domestic intervention scenarios*** | | | | | | | | | | | | |
| *Mtb* infection testing and treatment for new immigrants (A1) | 1.70 (1.49, 1.97) | 1.30 (1.09, 1.59) | 1.00 (0.74, 1.37) | 0.88 (0.55, 1.36) | 18 (17, 20) | 26 (24, 27) | 30 (29, 31) | 31 (30, 32) | 29 (24, 33) | 46 (38, 52) | 59 (45, 68) | 63 (44, 76) |
| Improved *Mtb* infection testing and treatment in U.S. risk populations (A2) | 1.72 (1.47, 2.02) | 1.29 (1.06, 1.63) | 0.99 (0.71, 1.40) | 0.88 (0.53, 1.41) | 18 (14, 20) | 26 (22, 30) | 31 (26, 34) | 32 (27, 35) | 29 (22, 34) | 46 (36, 54) | 59 (44, 70) | 63 (42, 77) |
| Improved TB case detection (A3) | 1.92 (1.68, 2.25) | 1.62 (1.33, 1.99) | 1.33 (0.98, 1.84) | 1.20 (0.74, 1.86) | 7 (5, 9) | 7 (6, 9) | 7 (5, 9) | 7 (5, 8) | 20 (13, 25) | 33 (22, 41) | 44 (27, 58) | 50 (23, 68) |
| Improved TB disease treatment outcomes (A4) | 1.99 (1.74, 2.32) | 1.68 (1.40, 2.05) | 1.38 (1.01, 1.89) | 1.24 (0.77, 1.91) | 4 (3, 4) | 4 (3, 4) | 4 (3, 4) | 4 (3, 4) | 17 (10, 22) | 30 (19, 39) | 43 (24, 56) | 48 (22, 67) |
| All domestic interventions (A5) | 1.23 (1.06, 1.45) | 0.85 (0.69, 1.07) | 0.63 (0.44, 0.89) | 0.55 (0.33, 0.88) | 41 (38, 43) | 52 (49, 54) | 56 (54, 59) | 57 (55, 60) | 49 (44, 53) | 65 (58, 70) | 74 (65, 81) | 77 (64, 86) |
| ***International intervention scenarios*** | | | | | | | | | | | | |
| Scale-up of current technology (B1) | 1.81 (1.59, 2.10) | 1.45 (1.21, 1.77) | 1.03 (0.77, 1.39) | 0.74 (0.47, 1.13) | 12 (9, 16) | 17 (13, 22) | 28 (25, 33) | 42 (39, 46) | 24 (18, 29) | 40 (31, 47) | 57 (44, 67) | 69 (53, 80) |
| Scale-up of current and new technology (B2) | 1.69 (1.47, 1.96) | 1.35 (1.12, 1.65) | 0.91 (0.68, 1.23) | 0.61 (0.39, 0.94) | 19 (14, 24) | 22 (18, 29) | 36 (33, 41) | 53 (50, 56) | 30 (24, 35) | 44 (35, 51) | 62 (50, 71) | 75 (62, 83) |
| Scale-up of current and new tech., and TB vaccine from 2030 (B3) | 1.60 (1.40, 1.86) | 1.08 (0.89, 1.32) | 0.55 (0.41, 0.74) | 0.30 (0.20, 0.48) | 23 (18, 29) | 38 (33, 44) | 62 (58, 67) | 76 (72, 80) | 33 (27, 39) | 55 (48, 61) | 77 (71, 82) | 87 (81, 91) |
| ***Combined international and domestic intervention scenarios*** | | | | | | | | | | | | |
| All interventions, excluding new TB vaccine (C1) | 1.00 (0.86, 1.20) | 0.63 (0.51, 0.79) | 0.37 (0.27, 0.52) | 0.24 (0.15, 0.39) | 52 (47, 56) | 64 (60, 67) | 74 (72, 76) | 81 (79, 83) | 58 (53, 62) | 74 (69, 77) | 84 (79, 88) | 90 (84, 94) |
| All interventions, with new TB vaccine internationally (C2) | 0.94 (0.81, 1.14) | 0.49 (0.40, 0.63) | 0.21 (0.16, 0.31) | 0.12 (0.07, 0.20) | 54 (50, 59) | 72 (69, 75) | 85 (83, 87) | 91 (89, 92) | 60 (56, 65) | 80 (75, 83) | 91 (88, 93) | 95 (92, 97) |

**Table S3: Projected future United States TB incidence rates, percentage reductions in incidence compared to base-case in same year, and percentage reductions in incidence rates compared to 2024 for each intervention scenario in selected years.** Values in parentheses represent 95% uncertainty intervals.

| | *Mtb* infections averted (%) | TB cases averted (%) | TB deaths averted (%) | Life-years saved (%) | QALYs saved (%) | Value of TB incidence reductions (%) |
|---|---|---|---|---|---|---|
| ***Domestic intervention scenarios*** | | | | | | |
| *Mtb* infection testing and treatment for new immigrants (A1) | 17.0 (14.9, 18.9) | 17.3 (15.8, 18.7) | 14.8 (13.2, 16.5) | 16.3 (14.7, 17.8) | 16.4 (14.8, 17.9) | 14.8 (13.3, 16.3) |
| Improved *Mtb* infection testing and treatment in U.S. risk populations (A2) | 16.5 (13.9, 18.8) | 16.7 (13.7, 19.2) | 17.7 (14.6, 20.1) | 16.7 (13.7, 19.1) | 16.7 (13.7, 19.1) | 15.5 (12.7, 17.8) |
| Improved TB case detection (A3) | 46.7 (44.6, 49.4) | 5.7 (4.2, 7.0) | 21.8 (20.2, 23.3) | 23.2 (21.1, 24.8) | 23.5 (21.6, 25.0) | 21.5 (19.6, 23.0) |
| Improved TB disease treatment outcomes (A4) | 3.6 (3.0, 4.4) | 3.6 (3.2, 3.9) | 6.4 (5.6, 7.0) | 7.2 (6.3, 7.9) | 7.0 (6.1, 7.6) | 6.8 (5.9, 7.4) |
| All domestic interventions (A5) | 63.4 (61.8, 65.0) | 37.2 (34.8, 39.3) | 48.7 (46.3, 50.6) | 50.1 (48.0, 51.9) | 50.2 (48.1, 52.0) | 47.1 (44.9, 49.0) |
| ***International intervention scenarios*** | | | | | | |
| Scale-up of current technology (B1) | 13.6 (10.6, 18.0) | 11.6 (8.9, 15.0) | 8.9 (6.1, 12.5) | 10.4 (7.8, 13.9) | 10.6 (7.9, 14.0) | 9.4 (6.9, 12.6) |
| Scale-up of current and new technology (B2) | 19.6 (15.4, 25.7) | 17.0 (13.3, 21.8) | 13.1 (9.0, 18.1) | 15.3 (11.6, 20.1) | 15.5 (11.9, 20.3) | 14.1 (10.4, 18.6) |
| Scale-up of current and new technology, and TB vaccine from 2030 (B3) | 25.1 (20.4, 31.2) | 22.9 (19.0, 27.8) | 17.4 (12.9, 22.6) | 20.7 (16.6, 25.9) | 20.9 (16.8, 26.1) | 18.5 (14.5, 23.4) |
| ***Combined international and domestic intervention scenarios*** | | | | | | |
| All interventions, excluding new TB vaccine (C1) | 72.1 (70.1, 73.9) | 47.3 (43.5, 51.1) | 54.8 (51.2, 58.0) | 57.2 (53.8, 60.0) | 57.4 (54.0, 60.2) | 53.8 (50.4, 56.7) |
| All interventions, with new TB vaccine internationally (C2) | 73.8 (71.8, 75.8) | 50.6 (46.7, 54.5) | 56.6 (52.9, 59.9) | 59.5 (56.1, 62.4) | 59.7 (56.3, 62.5) | 55.7 (52.3, 58.7) |

**Table S4: Estimated impact of base-case and intervention scenarios on cumulative health and economic outcomes, United States, 2024-2050 (percentage reduction compared to base-case).** QALYs = quality-adjusted life years, *Mtb* = *Mycobacterium tuberculosis*. Values in parentheses represent 95% uncertainty intervals.

| | TB treatment costs averted (USD mil.) | Averted productivity losses (USD mil.) | | Societal value of averted TB mortality (USD mil.) | Total economic value of TB incidence reductions (USD mil.)* |
|---|---|---|---|---|---|
| | | From TB treatment | From TB deaths | | |
| ***Domestic intervention scenarios*** | | | | | |
| *Mtb* infection testing and treatment for new immigrants (A1) | 591 (498, 719) | 65 (54, 79) | 1,779 (1,335, 2,308) | 17,450 (8,166, 29,055) | 18,151 (8,828, 29,822) |
| Improved *Mtb* infection testing and treatment in U.S. risk populations (A2) | 551 (446, 676) | 58 (46, 71) | 1,668 (1,235, 2,146) | 18,451 (8,216, 30,120) | 19,056 (8,771, 30,796) |
| Improved TB case detection (A3) | 97 (36, 170) | 11 (5, 17) | 2,607 (2,011, 3,240) | 26,323 (12,228, 42,733) | 26,422 (12,365, 42,820) |
| Improved TB disease treatment outcomes (A4) | 132 (112, 159) | 14 (12, 17) | 842 (641, 1,036) | 8,165 (3,796, 13,329) | 8,323 (3,951, 13,473) |
| All domestic interventions (A5) | 1,206 (1,011, 1,439) | 129 (109, 154) | 5,495 (4,284, 6,820) | 56,427 (26,235, 91,368) | 57,746 (27,605, 93,020) |
| ***International intervention scenarios*** | | | | | |
| Scale-up of current technology (B1) | 400 (292, 537) | 42 (31, 56) | 1,174 (786, 1,639) | 11,108 (5,023, 19,655) | 11,566 (5,469, 20,284) |
| Scale-up of current and new technology (B2) | 599 (445, 796) | 62 (46, 82) | 1,754 (1,187, 2,433) | 16,577 (7,495, 29,458) | 17,243 (8,144, 30,168) |
| Scale-up of current and new technology, and TB vaccine from 2030 (B3) | 783 (613, 1,010) | 85 (67, 108) | 2,343 (1,666, 3,158) | 21,893 (10,016, 37,837) | 22,739 (10,883, 38,957) |
| ***Combined international and domestic intervention scenarios*** | | | | | |
| All interventions, excluding new TB vaccine (C1) | 1,581 (1,321, 1,915) | 168 (140, 202) | 6,286 (4,881, 7,853) | 64,111 (29,814, 105,769) | 65,902 (31,594, 107,638) |
| All interventions, with new TB vaccine internationally (C2) | 1,685 (1,410, 2,031) | 180 (151, 217) | 6,540 (5,070, 8,178) | 66,365 (30,843, 109,460) | 68,138 (32,741, 111,804) |

**Table S5: Estimated impact of intervention scenarios on disaggregated economic outcomes, United States, 2024-2050.** USD = U.S. dollars. * The total economic value of TB incidence reductions calculated as the sum of other columns excluding the productivity losses associated with TB mortality. This is done to avoid double-counting, as productivity losses due to premature mortality are included in the societal value of averted TB deaths.

| Scenario | Year pre-elimination target met (incidence <1.0 per 100,000) | Year elimination target met (incidence <0.1 per 100,000) |
|---|---|---|
| Base-case scenario | >2099 (2079, >2099) | >2099 (>2099, >2099) |
| ***Domestic intervention scenarios*** | | |
| *Mtb* infection testing and treatment for new immigrants (A1) | 2075 (2055, >2099) | >2099 (>2099, >2099) |
| Improved *Mtb* infection testing and treatment in U.S. risk populations (A2) | 2074 (2052, >2099) | >2099 (>2099, >2099) |
| Improved TB case detection (A3) | >2099 (2073, >2099) | >2099 (>2099, >2099) |
| Improved TB disease treatment outcomes (A4) | >2099 (2076, >2099) | >2099 (>2099, >2099) |
| All domestic interventions (A5) | 2042 (2036, 2057) | >2099 (>2099, >2099) |
| ***International intervention scenarios*** | | |
| Scale-up of current technology (B1) | 2077 (2060, >2099) | >2099 (>2099, >2099) |
| Scale-up of current and new technology (B2) | 2069 (2056, 2092) | >2099 (>2099, >2099) |
| Scale-up of current and new technology, and TB vaccine from 2030 (B3) | 2052 (2046, 2061) | >2099 (>2099, >2099) |
| ***Combined international and domestic intervention scenarios*** | | |
| All interventions, excluding new TB vaccine (C1) | 2034 (2031, 2040) | >2099 (>2099, >2099) |
| All interventions, with new TB vaccine internationally (C2) | 2033 (2031, 2037) | >2099 (2088, >2099) |

**Table S6: Year in which pre-elimination and elimination targets met for total U.S population, for base-case and intervention scenarios.** Point estimates represent median year in which target met. Values in parentheses represent 95% uncertainty intervals. Under a sensitivity analysis in which the coverage and effectiveness of domestic interventions was adjusted to match the targets of the End TB Strategy as applied in international scenarios, U.S. TB incidence rates declined rapidly, with pre-elimination and elimination targets reached in 2026 (2025, 2026), and 2073 (2062, >2099) respectively.

| Scenario | TB incidence rate in 2050 (per 100,000) | | | Incidence rate relative to value estimated in main analysis (%) | |
|---|---|---|---|---|---|
| | Main analysis | Future migration rate 50% lower | Future migration rate 50% higher | Future migration rate 50% lower | Future migration rate 50% higher |
| Base-case scenario (continuation of current trends) | 1.75 (1.46, 2.13) | 1.08 (0.90, 1.32) | 2.35 (1.95, 2.88) | -38.1 (-39.9, -36.3) | 34.8 (33.3, 36.3) |
| ***Domestic intervention scenarios*** | | | | | |
| *Mtb* infection testing and treatment for new immigrants (A1) | 1.30 (1.09, 1.59) | 0.84 (0.71, 1.03) | 1.71 (1.44, 2.09) | -35.1 (-37.4, -32.8) | 32.0 (30.2, 34.0) |
| Improved *Mtb* infection testing and treatment in U.S. risk populations (A2) | 1.29 (1.06, 1.63) | 0.77 (0.63, 0.98) | 1.76 (1.44, 2.22) | -40.0 (-41.4, -38.4) | 36.5 (35.2, 37.7) |
| Improved TB case detection (A3) | 1.62 (1.33, 1.99) | 0.99 (0.82, 1.22) | 2.19 (1.81, 2.70) | -39.1 (-40.6, -37.6) | 35.7 (34.4, 37.1) |
| Improved TB disease treatment outcomes (A4) | 1.68 (1.40, 2.05) | 1.04 (0.87, 1.27) | 2.26 (1.87, 2.77) | -38.3 (-40.0, -36.5) | 34.9 (33.5, 36.4) |
| All domestic interventions (A5) | 0.85 (0.69, 1.07) | 0.52 (0.43, 0.66) | 1.14 (0.94, 1.45) | -38.2 (-39.9, -36.5) | 34.9 (33.4, 36.4) |
| ***International intervention scenarios*** | | | | | |
| Scale-up of current technology (B1) | 1.45 (1.21, 1.77) | 0.92 (0.77, 1.12) | 1.94 (1.61, 2.35) | -36.5 (-38.5, -34.6) | 33.3 (31.7, 35.0) |
| Scale-up of current and new technology (B2) | 1.35 (1.12, 1.65) | 0.87 (0.73, 1.06) | 1.79 (1.49, 2.18) | -35.8 (-37.8, -33.8) | 32.7 (31.0, 34.4) |
| Scale-up of current and new tech., and TB vaccine from 2030 (B3) | 1.08 (0.89, 1.32) | 0.72 (0.60, 0.89) | 1.40 (1.16, 1.72) | -32.9 (-35.2, -30.6) | 30.0 (28.0, 32.0) |
| ***Combined international and domestic intervention scenarios*** | | | | | |
| All interventions, excluding new TB vaccine (C1) | 0.63 (0.51, 0.79) | 0.41 (0.33, 0.51) | 0.83 (0.68, 1.05) | -35.5 (-37.5, -33.7) | 32.4 (30.9, 34.0) |
| All interventions, with new TB vaccine internationally (C2) | 0.49 (0.40, 0.63) | 0.33 (0.27, 0.43) | 0.63 (0.51, 0.81) | -32.1 (-34.4, -30.1) | 29.3 (27.5, 31.3) |

**Table S7: Results of sensitivity analyses with alternative values for selected parameters.** Values in parentheses represent 95% uncertainty intervals.

| Model input | Partial rank correlation coefficient | | |
|---|---|---|---|
| | Point estimate | Lower uncertainty interval | Upper uncertainty interval |
| Annual trends in future migration growth post-2024 | 0.70 | 0.66 | 0.75 |
| Rate of progression from established *Mtb* infection to TB disease | 0.56 | 0.51 | 0.62 |
| Rate of testing for *Mtb* infection within recommended groups within the United States | -0.48 | -0.53 | -0.42 |
| Immigration volume in 2024 | 0.35 | 0.29 | 0.41 |
| TB disease prevalence among migrants entering the United States | 0.33 | 0.27 | 0.39 |
| Rate ratio of progression from established *Mtb* infection to TB disease for non-U.S.-born vs. U.S.-born | 0.22 | 0.16 | 0.28 |
| *Mtb* infection prevalence among migrants entering the United States | 0.16 | 0.10 | 0.22 |
| Annual rate of decline in TB progression rate for individuals with established *Mtb* infection | -0.16 | -0.22 | -0.10 |

**Table S8: Partial rank correlation coefficients for model projection of U.S. TB incidence rate in 2050 under base-case scenario (parameters with greatest absolute coefficients shown).**

### References

1. Menzies NA, Cohen T, Hill AN, Yaesoubi R, Galer K, Wolf E, Marks SM, Salomon JA. Prospects for Tuberculosis Elimination in the United States: Results of a Transmission Dynamic Model. *Am J Epidemiol* 2018; 187: 2011-2020.

2. Sterling TR, Bethel J, Goldberg S, Weinfurter P, Yun L, Horsburgh CR. The scope and impact of treatment of latent tuberculosis infection in the United States and Canada. *Am J Respir Crit Care Med* 2006; 173: 927-931.

3. US Preventive Services Task Force. Screening for Latent Tuberculosis Infection in Adults: US Preventive Services Task Force Recommendation Statement. *JAMA* 2023; 329: 1487-1494.

4. U.S. Centers for Disease Control and Prevention. Latent tuberculosis infection: A guide for primary health care providers [https://www.cdc.gov/tb/publications/ltbi/pdf/LTBIbooklet508.pdf]. Atlanta GA, USA: U.S. Department of Health and Human Services; 2020.

5. Stout JE, Wu Y, Ho CS, Pettit AC, Feng PJ, Katz DJ, Ghosh S, Venkatappa T, Luo R. Evaluating latent tuberculosis infection diagnostics using latent class analysis. *Thorax* 2018; 73: 1062-1070.

6. U.S. Centers for Disease Control and Prevention. 2021 State and City TB Report. Atlanta GA: Division of Tuberculosis Elimination, National Center for HIV, Viral Hepatitis, STD, and TB Prevention, U.S. Centers for Disease Control and Prevention; 2021.

7. Menzies D, Adjobimey M, Ruslami R, Trajman A, Sow O, Kim H, Obeng Baah J, Marks GB, Long R, Hoeppner V, Elwood K, Al-Jahdali H, Gninafon M, Apriani L, Koesoemadinata RC, Kritski A, Rolla V, Bah B, Camara A, Boakye I, Cook VJ, Goldberg H, Valiquette C, Hornby K, Dion MJ, Li PZ, Hill PC, Schwartzman K, Benedetti A. Four Months of Rifampin or Nine Months of Isoniazid for Latent Tuberculosis in Adults. *N Engl J Med* 2018; 379: 440-453.

8. Sterling TR, Villarino ME, Borisov AS, Shang N, Gordin F, Bliven-Sizemore E, Hackman J, Hamilton CD, Menzies D, Kerrigan A, Weis SE, Weiner M, Wing D, Conde MB, Bozeman L, Horsburgh CR, Jr., Chaisson RE. Three months of rifapentine and isoniazid for latent tuberculosis infection. *N Engl J Med* 2011; 365: 2155-2166.

9. International Union Against Tuberculosis Committee on Prophylaxis. Efficacy of various durations of isoniazid preventive therapy for tuberculosis: five years of follow-up in the IUAT trial. *Bull World Health Organ* 1982; 60: 555-564.

10. Feng PI, Horne DJ, Wortham JM, Katz DJ. Trends in tuberculosis clinicians' adoption of short-course regimens for latent tuberculosis infection. *J Clin Tuberc Other Mycobact Dis* 2023; 33: 100382.

11. Sandul AL, Nwana N, Holcombe JM, Lobato MN, Marks S, Webb R, Wang SH, Stewart B, Griffin P, Hunt G, Shah N, Marco A, Patil N, Mukasa L, Moro RN, Jereb J, Mase S, Chorba T, Bamrah-Morris S, Ho CS. High Rate of Treatment Completion in Program Settings With 12-Dose Weekly Isoniazid and Rifapentine for Latent Mycobacterium tuberculosis Infection. *Clin Infect Dis* 2017; 65: 1085-1093.

12. U. S. Centers for Disease Control and Prevention. Tuberculosis Technical Instructions for Panel Physicians. Atlanta, GA; 2019.

13. Khan A, Phares CR, Phuong HL, Trinh DTK, Phan H, Merrifield C, Le PTH, Lien QTK, Lan SN, Thoa PTK, Thu LTM, Tran T, Tran C, Platt L, Maloney SA, Nhung NV, Nahid P, Oeltmann JE. Overseas Treatment of Latent Tuberculosis Infection in US-Bound Immigrants. *Emerg Infect Dis* 2022; 28: 582-590.

14. U.S. Centers for Disease Control and Prevention. Reported Tuberculosis in the United States, 2021. Atlanta, GA; 2022.

15. Stop TB Partnership. The global plan to end TB 2023-2030. Geneva: Stop TB Partnership; 2022.

16. WHO Global TB Programme. WHO Global TB Database [http://www.who.int/tb/country/data/download/en/]. Geveva Switzerland: WHO Global TB Programme; 2023.

17. World Health Organization. The End TB Strategy 2015. Geneva, Switzerland: World Health Organization; 2016.

18. Houben RM, Lalli M, Sumner T, Hamilton M, Pedrazzoli D, Bonsu F, Hippner P, Pillay Y, Kimerling M, Ahmedov S, Pretorius C, White RG. TIME Impact - a new user-friendly

tuberculosis (TB) model to inform TB policy decisions. *BMC Med* 2016; 14: 56. doi: 10.1186/s12916-12016-10608-12914.

19. World Health Organization. Global TB Report 2022. Geneva; 2022.

20. Menzies NA, Bellerose M, Testa C, Swartwood NA, Malyuta Y, Cohen T, Marks SM, Hill AN, Date AA, Maloney SA, Bowden SE, Grills AW, Salomon JA. Impact of Effective Global Tuberculosis Control on Health and Economic Outcomes in the United States. *Am J Respir Crit Care Med* 2020; 202: 1567-1575.

21. Arias E, Xu J. United States Life Tables, 2019. *National Vital Statistics Reports* 2022; 70.

22. Castro KG, Marks SM, Chen MP, Hill AN, Becerra JE, Miramontes R, Winston CA, Navin TR, Pratt RH, Young KH, LoBue PA. Estimating tuberculosis cases and their economic costs averted in the United States over the past two decades. *Int J Tuberc Lung Dis* 2016; 20: 926-933.

23. Winston CA, Marks SM, Carr W. Estimated Costs of 4-Month Pulmonary Tuberculosis Treatment Regimen, United States. *Emerg Infect Dis* 2023; 29: 2102-2104.

24. Dunn A, Grosse SD, Zuvekas SH. Adjusting Health Expenditures for Inflation: A Review of Measures for Health Services Research in the United States. *Health Services Research* 2018; 53: 175-196.

25. Sanders GD, Neumann PJ, Basu A, Brock DW, Feeny D, Krahn M, Kuntz KM, Meltzer DO, Owens DK, Prosser LA, Salomon JA, Sculpher MJ, Trikalinos TA, Russell LB, Siegel JE, Ganiats TG. Recommendations for Conduct, Methodological Practices, and Reporting of Cost-effectiveness Analyses: Second Panel on Cost-Effectiveness in Health and Medicine. *JAMA* 2016; 316: 1093-1103. doi: 1010.1001/jama.2016.12195.

26. Grosse SD, Krueger KV, Pike J. Estimated annual and lifetime labor productivity in the United States, 2016: implications for economic evaluations. *Journal Med Econ* 2019; 22: 501-508.

27. Bureau of Labor Statistics. CUUR0000SA0: CPI for All Urban Consumers (CPI-U) [https://data.bls.gov/timeseries/CUUR0000SA0?years_option=all_years]. Washington DC, USA: Bureau of Labor Statistics; 2023.

28. Bureau of Labor Statistics. LEU0252881600: Constant (1982-84) dollar adjusted to CPI-U-Median usual weekly earnings, Employed full time, Wage and salary workers [https://data.bls.gov/cgi-bin/surveymost]. Washington DC, USA: Bureau of Labor Statistics; 2023.

29. Taylor Z, Marks SM, Rios Burrows NM, Weis SE, Stricof RL, Miller B. Causes and costs of hospitalization of TB patients in the United States *Int J Tuberc Lung Dis* 2000; 4: 931-939.

30. Shepardson D, Marks SM, Chesson H, Kerrigan A, Holland DP, Scott N, Tian X, Borisov AS, Shang N, Heilig CM, Sterling TR, Villarino ME, Mac Kenzie WR. Cost-effectiveness of a 12-dose regimen for treating latent tuberculous infection in the United States. *Int J Tuberc Lung Dis* 2013; 17: 1531-1537.

31. Congressional Budget Office. The 2021 Long-Term Budget Outlook. Washington, DC: Congressional Budget Office; 2021.

32. Robinson LA, Hammitt JK, Baxter JR. Guidelines For Regulatory Impact Analysis [retrieved from https://aspe.hhs.gov/pdf-report/guidelines-regulatory-impact-analysis, June 3 2019]. Washington DC: U.S. Department of Health and Human Services; 2016.

33. Robinson LA, Hammitt JK. Valuing Reductions in Fatal Illness Risks: Implications of Recent Research. *Health Econ* 2016; 25: 1039-1052.

34. ASPE. HHS Guidelines for Regulatory Impact Analysis Appendix D: Updating Value per Statistical Life (VSL) Estimates for Inflation and Changes in Real Income Washington DC: HHS Office of the Assistant Secretary for Planning and Evaluation (ASPE); 2021.

35. Houben RM, Dodd PJ. The Global Burden of Latent Tuberculosis Infection: A Re-estimation Using Mathematical Modelling. *PLoS Med* 2016; 13: e1002152. doi: 1002110.1001371/journal.pmed.1002152. eCollection 1002016 Oct.
